# Investigating Primary Care Indications to Improve the Quality of Electronic Health Record Data in Target Trial Emulation for Dementia

**DOI:** 10.1101/2025.04.08.25325485

**Authors:** Max Sunog, Colin Magdamo, Marie-Laure Charpignon, Mark Albers

**Affiliations:** Massachusetts General Hospital, US; Harvard Medical School, US; Institute for Data, Systems, and Society, Massachusetts Institute of Technology & Computational Health Informatics Program, Boston Children’s Hospital & Harvard Medical School, US; Massachusetts General Hospital & Harvard Medical School, US

## Abstract

Missing data, inaccuracies in medication lists, and recording delays in electronic health records (EHR) are major limitations for target trial emulation (TTE), which uses EHR data to retrospectively emulate a clinical trial. EHR-based TTE relies on recorded data that proxy actual drug exposures and outcomes. While prior work has proposed various methods to improve EHR data quality, here we investigate the underutilized consideration that encounters with a primary care provider (PCP) may result in more accurate data in the EHR. Patients with a PCP within the EHR network being studied tend to have more encounters overall and a greater proportion of the types of encounters that yield comprehensive and up-to-date records. By contrasting data for patients with and without a PCP in the considered EHR network, we demonstrate how PCP status affects EHR data quality. Through a case study, we then empirically examine the impact on TTE of including a PCP status feature either in the propensity score and outcome models or as an eligibility criterion for cohort selection, versus ignoring it. Specifically, we compare the estimated effects of two first-line antidiabetic drug classes on the onset of Alzheimer’s Disease and Related Dementias. We find that the estimated treatment effect is sensitive to the consideration of PCP status, particularly when used as an eligibility criterion. Our work suggests that further researching the role of PCP status may improve the design of pragmatic trials.

**Data and Code Availability:** The study uses EHR data from the Research Patient Data Registry (Nalichowski et al., 2007), social vulnerability index (SVI) data from the Agency for Toxic Substances and Disease Registry (https://www.atsdr.cdc.gov/placeandhealth/svi), and Massachusetts death records from the Registry of Vital Records and Statistics. Because the data contain patient information, they cannot be made available.

**Institutional Review Board (IRB):** This research was performed under MGB IRB approval (protocol 2023P000604).

## 1. Introduction

The widespread adoption of electronic health records (EHR) to collect healthcare information has generated large stores of structured data. Through the target trial emulation (TTE) framework, these data can be used to emulate otherwise infeasible studies when traditional randomized controlled trials (RCT) are prohibited due to ethical reasons, inability to recruit participants, or the extensive duration required for the trial. For example, RCTs examining the onset of Alzheimer’s Disease and Related Dementias (ADRD) are infeasible because of the extended preclinical phase of the disease, which can last fifteen years or more (Bateman et al., 2012). However, EHR-based TTE makes testing drug repurposing hypotheses related to ADRD onset possible; one such study found a protective effect of initiating the antidiabetic drug metformin vs. sulfonylureas on incident ADRD (Charpignon et al., 2022).

In EHR-based TTE, patients are *enrolled* based on records of treatment initiation. Further, their followup and censoring times are *derived* from records of an outcome or last visit. Because these are indirect observations, the estimated risk of disease (e.g., ADRD onset) is that of an *outcome being recorded* (e.g., having an ADRD diagnosis), given the *recorded treatment(s)*. Investigators often implicitly assume that the estimated risk reflects the risk that relates the actual treatment assignment to the actual outcome; for instance, the result that patients with an initial prescription record of metformin (versus sulfonylureas) have a lower risk of recorded ADRD onset suggests that patients who actually initiate metformin versus sulfonylureas similarly have a lower risk of developing ADRD.

Under the premise that recorded data reflect actual treatments and outcomes, studies have addressed some methodological limitations. For example, emulated trials can reduce confounding through inverse propensity of treatment weighting (IPTW), a process in which a model is trained to estimate the probability that a patient receives a certain treatment (Rosenbaum and Rubin, 1983). Using this model, scores are estimated for each patient’s propensity of receiving their treatment, and they are assigned case weights for outcome modeling based inversely related to those scores. This method balances the treatment arms for measured confounders, thereby removing their impact on the estimated effect of the treatment itself. Additionally, TTEs can account for competing risks to the primary outcome (e.g., death before developing ADRD). Without considering the competing risk of death, patients who die are treated like patients who are lost to follow-up even though those who die cannot eventually develop the primary outcome, while the latter can. In trials where treatment arms have different mortality rates, treating death as a competing risk can reduce the bias of risk estimates for the primary outcome (Andersen et al., 1993).

While valuable, these methods do not address the strong assumption that EHR data accurately reflect events that occur in nature. In fact, they often rely heavily on the accuracy of recorded data; just as up-to-date death records are necessary to account for the competing risk of death, IPTW requires comprehensive records to appropriately define confounders. Therefore, accurate and timely records are needed to interpret EHR-based TTE findings, learn about the real-world effectiveness of candidate treatment strategies, and ultimately inform clinical decisions, such as switching a patient’s treatment to metformin in light of the drug’s estimated protective effect against ADRD. Unfortunately, missed or unrecorded diagnoses, inaccurate drug lists, and severe delays in diagnosis recording are common. As a result, EHR-based phenotyping often lacks sensitivity: a metareview of algorithms used to detect dementia in EHR data found that their sensitivity ranged from 8% to 79% when compared to expert clinical evaluation or chart review (Walling et al., 2023). While generally problematic, missingness in EHR data can even more detrimentally affect patients in minority groups or with lower socioeconomic status, as they are more likely to have missing records (Getzen et al., 2023).

To address this limitation, other approaches must be employed to improve EHR data quality. Principal factors associated with higher quality EHR data are 1) many recorded encounters, 2) types of procedures that inform thorough medical records (taking patient history, running regular screens, etc.), and 3) interactions with a provider who actively inputs records into the EHR (Verheij et al., 2018). Given these conditions, one under-utilized metric to identify patients with high-quality data is whether they see a Primary Care Physician (PCP) within the EHR’s healthcare system. At annual wellness visits, PCPs are likely to document a complete review and comprehensive history of their patients, and to update their EHR Sleath et al. (1999), and patients often regularly visit their PCP regardless of their health. These important visits are only recorded in the EHR used by the PCP, so in locations where patients have access to clinics within several EHR networks, the effect on a patient’s data quality in a specific EHR is dependent on their PCP practicing in that network. Therefore, we propose the use of carefully selected indications that a patient has an internal PCP encounter to improve records by 1) increasing the likelihood that patients in the cohort have enough visits to allow for a thorough history and follow-up during the study period, and 2) ensuring that these patients have the types of visits that materialize accurate medical records in the EHR. While prior work has utilized the total level of healthcare utilization in EHR-based TTE (Goldstein et al., 2016), this study adds a complementary method that crucially accounts for the types of encounters.

In this study, we 1) developed a definition of having an internal PCP by mining the EHR, 2) demonstrated that the quality of data recording in the EHR was higher among patients that have an internal PCP, and 3) explored the effects on TTE results of using internal PCP status as a feature in propensity score and outcome modeling, or as an eligibility criterion.

## 2. Methods

### 2.1. Identifying patients with an internal PCP

We identified three useful types of PCP indications present in our EHR system, the Research Patient Data Registry (RPDR): 1) procedure codes associated with primary care visits (e.g., annual wellness exams); 2) encounters under the ‘Primary Care’ service line, a categorization for visits performed by a PCP; and 3) encounters with ‘Annual Wellness Visit’ as the listed reason-for-visit (a code attached to encounters logged in Epic). Using these three types of, we defined a composite metric: a patient was assigned a positive PCP status if they had at least one such indication before treatment initiation, and a negative PCP status otherwise.

### 2.2 Evaluating the quality of baseline covariate data by PCP status

In our cohort of patients with type 2 diabetes (T2D), individuals who met the internal PCP definition had higher healthcare utilization and recorded prevalence rates of various diseases than those who did not (Table 1). Although the large disparity between PCP and no-PCP patients in the total number of visits before baseline owes in part to their longer medical history in the EHR network, the difference in the number of visits in the year before baseline reveals that PCP patients also have a higher rate of encounters.

**Table 1:** Summary statistics, comparing patients with (PCP) and without (No-PCP) a record indicative of an internal primary care provider prior to baseline. Covariates are measured at baseline, the date of the patient’s first prescription of metformin or a sulfonylurea. Outcomes of ADRD and death are prevalences after baseline. For age, social vulnerability scores (SVS), and visit metrics, means are reported with standard deviations. For sex, education (ed) levels, and covariate diagnoses (Dx), percentage values are reported.

| Feature | No-PCP | PCP |
| --- | --- | --- |
| Number of Patients | 37, 526 | 17, 112 |
| ADRD outcome | 5.7% | 5.0% |
| Death outcome | 21.1% | 9.8% |
| Age at Baseline | 66.6 (9.7) | 64.5 (8.9) |
| Sex Female | 49.9% | 53.4% |
| Ed. Secondary | 31.8% | 36.6% |
| Ed. College | 35.2% | 39.1% |
| Ed. Graduate | 7.5% | 8.6% |
| Ed. Missing | 25.5% | 15.7% |
| Socioeconomic SVS | 0.32 (.21) | 0.36 (.24) |
| Home Life SVS | 0.44 (.20) | 0.47 (.21) |
| Racial/Ethnic SVS | 0.41 (.21) | 0.48 (.23) |
| Housing SVS | 0.50 (.18) | 0.54 (.18) |
| Hypertension Dx | 56% | 81% |
| Stroke Dx | 4.1% | 6.5% |
| Cancer Dx | 29.8% | 38.7% |
| COPD Dx | 2.9% | 5.6% |
| Overweight Dx | 3.9% | 22.1% |
| Obesity Dx | 20.6% | 55.9% |
| CVD Dx | 17.0% | 25.6% |
| HbA1c Missing | 57.5% | 8.1% |
| HbA1c Reference | 3.6% | 7.0% |
| HbA1c Prediabetes | 11.8% | 29.3% |
| HbA1c Diabetes | 27.1% | 55.7% |
| BP Missing | 80.4% | 32.9% |
| BP Reference | 8.4% | 31.8% |
| BP Hypertension 1 | 4.6% | 17.8% |
| BP Hypertension 2 | 6.7% | 17.5% |
| Total Visits | 33.8 (50.3) | 128.2 (123.0) |
| Visits | 6.2 (11.1) | 17.3 (17.1) |
| Outpatient Visits | 27.8 (44.2) | 116.4 (115.3) |
| Outpatient Visits | 5.3 (10.2) | 16.6 (16.7) |
| Years in EHR | 15.9 (7.4) | 20.5 (7.7) |

When comparing the comorbidity distribution of our cohort with that of adults diagnosed with T2D in prior observational studies using well-phenotyped medical histories, we found a better alignment between the prevalence rates in PCP group and those reported in the literature; the no-PCP group had consistently lower prevalence rates. For instance, while Mamillapalli et al. (2019) found that 10%-20% of patients with T2D also suffered from chronic obstructive pulmonary disease (COPD), the PCP and no-PCP groups had prevalence rates of 5.6% and 2.9%, respectively. Similarly, 49.1% of T2D patients were found to have obesity (Nguyen et al., 2010); our PCP and no-PCP groups have obesity prevalence rates of 55.9% and 20.6%, respectively (Table 1). Although the populations captured in prior studies do not exactly match our cohort, the better alignment with the PCP group suggests that patients with an internal PCP have a more complete EHR than those without. In our real-world data applications, these covariates are used to balance treatment arms through IPTW, so their accuracy is critical to estimate causal effects with limited bias.

Although the lower diagnosis rates among patients without a PCP may be caused by missing records, they could also reflect actual population differences. This possibility raises a challenge; patients who receive a given diagnosis but have no corresponding record in the EHR are often indistinguishable from patients who did not receive that diagnosis. Fortunately, measurements such as HbA1c and blood pressure, as well as certain demographic characteristics such as educational attainment, can be specifically listed as missing (i.e. never captured in the EHR). In these cases, we can investigate whether differential rates are most likely caused by missingness or by population differences.

When restricting to patients without missing educational attainment records, the percentages of patients within each level were similar in the No-PCP vs. PCP group: 42.7% vs. 43.4% have secondary education, 47.2% vs. 46.4% have college education, and 10.1% vs. 10.2% have graduate education. Nonmissing HbA1c levels were less aligned, but similar: 8.5% vs. 7.6% had normal levels, 27.8% vs. 31.8% had prediabetic levels, and 63.8% vs. 60.5% have diabetic levels. Notably, more than half of No-PCP patients have no recorded HbA1c value prior to receiving an antidiabetic prescription, though HbA1c tests are typically ordered for the diagnosis and management of diabetes. With blood pressure, the differences were more apparent: 42.6% vs 47.4% had normal blood pressure, 23.4% vs. 26.5% had stage one hypertension, and 34.0% vs. 26.1% had stage two hypertension.

These comparisons cannot confirm whether the populations are actually aligned along these features, as the patients with missing values may be skewed differently (a particularly important consideration for blood pressure because 80.4% of No-PCP patients have no records). However, the much closer alignment between the PCP and No-PCP groups when restricting to patients with non-missing values does suggest that the disparities with respect to other features may largely owe to missingness.

### 2.3 Evaluating the quality of outcome ascertainment data

To investigate the disparity in mortality rate between PCP and no-PCP patients, we compared death records in the EHR with those provided by the Massachusetts (MA) department of public health, an accurate and complete source of information about deaths that occurred in the state in 2014-2024. In TTE applications, we use the state death registry to offset missingness caused by reporting delays affecting EHR data, which are common for deaths that occured within the last three years. Here, we restricted our evaluation to death events that occurred between 2014-2021 to avoid the effect of these delays. Using 5 features (first name, last name, birth date, sex, and zip code of residence), we matched patients in our cohort who had an MA zip code to records in the state death registry. Among the 632, 422 records present in the death registry, there was only a single collision on this combination of features. Thus, we considered any complete match with the EHR cohort highly likely to be a valid match.

Among the 3, 638 patients in our cohort with likely valid matches, 42.9% of no-PCP patients and 25.6% of PCP patients had no corresponding death record in the EHR. Given the strong matching criteria and comprehensiveness of the state death registry, these discrepancies mostly likely result from missing EHR death records. The pronounced dissimilarity between the PCP and No-PCP groups reinforces our hypothesis that PCP patients have fewer missing records in general. While the elevated missingness among No-PCP patients may have a variety of causes, our analysis suggests that PCP status is a strong proxy for the probability of having accurate death records in the EHR.

To evaluate PCP status against other healthcare utilization metrics that could be used to improve the quality of EHR data, we repeated the matching process using cohorts restricted to patients in the top quartile and decile of the number of visits they had prior to antidiabetic prescription (baseline). We considered five types of visits: outpatient, inpatient, specialist, emergency, and all. We found that using all visits generally led to the best data quality, so we chose this metric as the main comparator (results for other metrics are presented in Appendices G and H). We constructed cohorts using the 75^*th*^ (*V isits*_75_) and 90^*th*^ (*V isits*_90_) percentiles of this metric (calculated on the original, unfiltered cohort) as cutoffs, resulting in cohort sizes of 29,160 for *V isits*_75_ and 12,891 for *V isits*_90_.

In *V isits*_75_ and *V isits*_90_, 31.4% and 25.4% of patients with likely valid matches in the state registry were missing a corresponding EHR death record, respectively. In this experiment, *V isits*_90_ had marginally less missingness than the PCP cohort, but notably was 25% smaller. Overall, using the total number of visits as the primary healthcare utilization metric appears to reduce data missingness similarly to the application of the PCP criterion.

Despite having fewer missing death records, PCP patients have less than half the mortality rate (as calculated by the EHR records) of no-PCP patients. In fact, when MA death records are incorporated, PCP patients consistently have a much lower age-and-sex-specific mortality rate than no-PCP patients. By comparing these age-and-sex-specific mortality rates to those in the publicly available U.S. Census life tables for the entire MA population (Arias, 2022), we found that PCP patients aligned very closely with the published data, while the no-PCP patients at every age had roughly twice the death rate (Figure 2). To quantify the alignment, we compared the mean squared error (MSE) of the recorded mortality rates, weighted by the number of person-years of recorded time at that age among the population. The PCP population mortality rates had an MSE of 1.3 deaths per 1,000 person-years vs. 8.1 deaths per 1,000 person-years for the No-PCP group.

**Figure 1:**
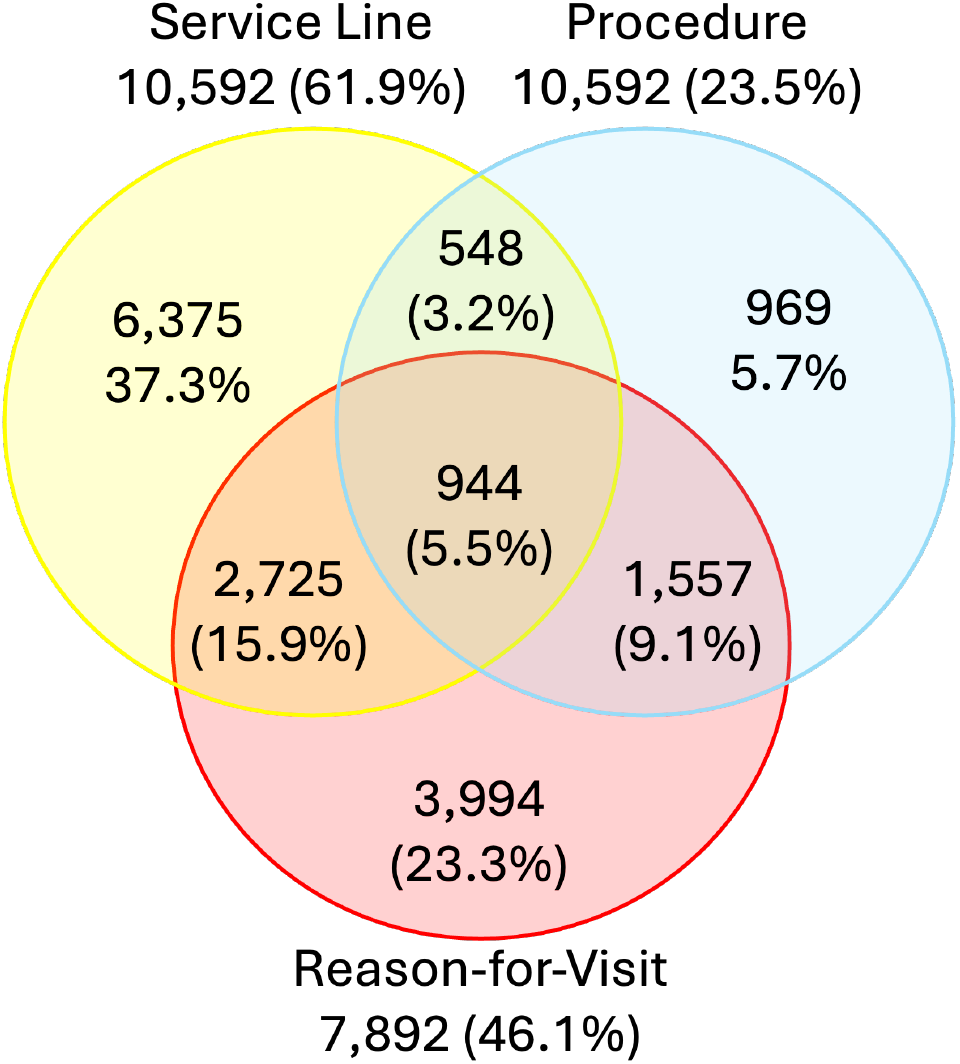
Venn diagram summarizing the number of patients by type of PCP indication (i.e., procedure, service line, and reason-for-visit) present in their EHR prior to their first antidiabetic prescription. Percentages are calculated out of the 17,112 patients with any PCP indication prior to their first antidiabetic prescription. Percentages of patients with each specific indication out of those with any indication are provided under each label. The percentage of patients with each specific combination of indications are provided in the circles.

**Figure 2:**
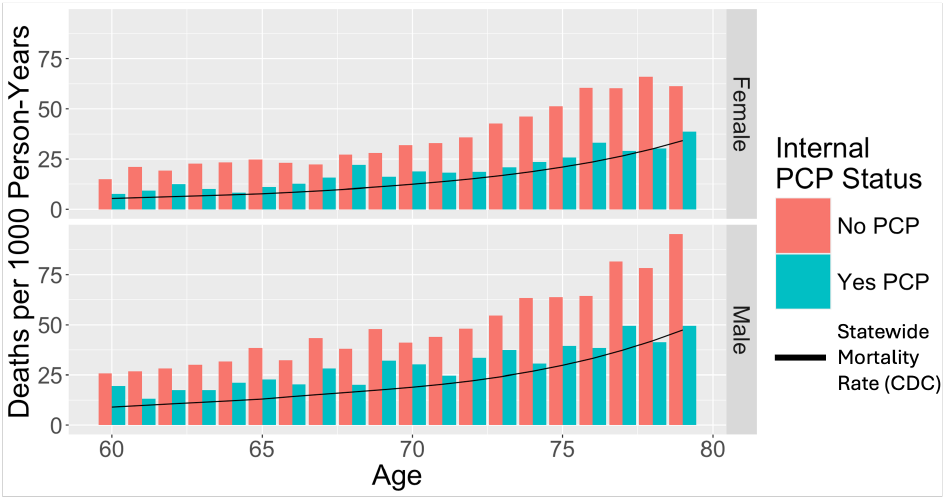
Age-specific mortality rate per 1,000 person-years among PCP vs. no-PCP patients, stratified by sex. Sex-specific trend lines in black correspond to official US census mortality rates in MA (Arias, 2022).

The MA death records should not be affected by PCP status or healthcare utilization, so the disparity is not a result of data quality difference. Instead, it demonstrates an actual elevated mortality rate among patients with hospital records but no internal PCP, which is expected because patients receiving tertiary care are likely have more severe illness relative to those receiving primary care. Therefore, a potential advantage of selecting only patients with an internal PCP is that their overall health is more representative of the general population.

In *V isits*_75_, the MSE was 4.9 deaths per 1,000 person-years, a significantly weaker alignment than in the PCP cohort. Notably, in *V isits*_90_, the MSE was 6.2 deaths per 1,000 person-years, revealing worse alignment with stricter filtering. The error in both cohorts are from consistently elevated mortality rates, which may be caused by the selection on patients who frequently visit the hospital, and therefore are likely to have more severe illness (Figure 3). This illustrates a disadvantage of using number of visits as a healthcare metric, as it can potentially induce a selection bias on particularly ill patients.

**Figure 3:**
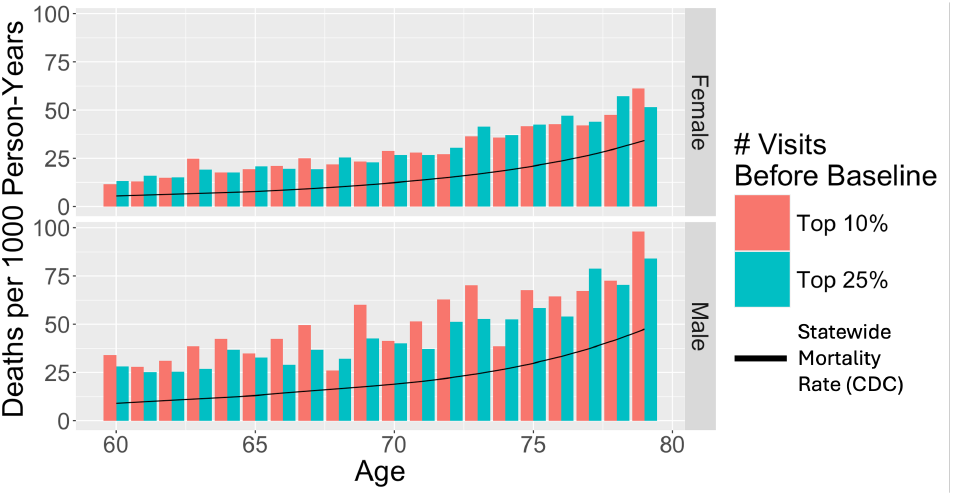
Age-specific mortality rate per 1,000 person-years among patients in the top decile vs. top quartile, based on their number of visits prior to antidiabetic prescription (baseline), stratified by sex. Sex-specific trend lines in black correspond to official US census mortality rates in MA (Arias, 2022).

In addition to all-cause mortality outcomes, we compared the PCP and no-PCP patient groups with respect to their ADRD outcomes. To that end, we used as our reference a study reporting age-specific incident ADRD diagnosis rate (as defined by the CMS Chronic Condition Warehouse algorithm) per person-year estimated from Medicare claims data for over 8 million patients across the US (Olfson et al., 2021). Our cohort differs from the overall Medicare population both because every patient in the cohort is diabetic, and in demographics such as race, ethnicity, and social vulnerability. Still, we expect that a replication of their methodology using the same set of diagnosis codes should result in similar incidence rates, provided that detection, diagnosis, and recording practices aligned.

For ages up to 75, both of the PCP and no-PCP groups aligned well with the reference study. Above 75 – when ADRD incidence is most common and alignment most critical – the PCP group had incidence rates similar to the reference study, while the no-PCP group consistently had roughly 40% lower incidence rates than the reference (Figure 4), suggesting that ADRD detection and diagnosis recording is improved among PCP patients, and that no-PCP patients are more likely to have missing ADRD records. The PCP cohort had a weighted MSE of 1.0 ADRD diagnoses per 1,000 person-years, compared to 8.7 for the no-PCP cohort, 1.5 for *V isits*_75_, and 1.4 for *V isits*_90_. Although this alignment can be impacted by population differences, ADRD diagnosis coding has particularly high missingness in the EHR, so data quality plays a large role. In this case, the alternative healthcare utilization metrics provide nearly as much alignment to the PCP criterion.

**Figure 4:**
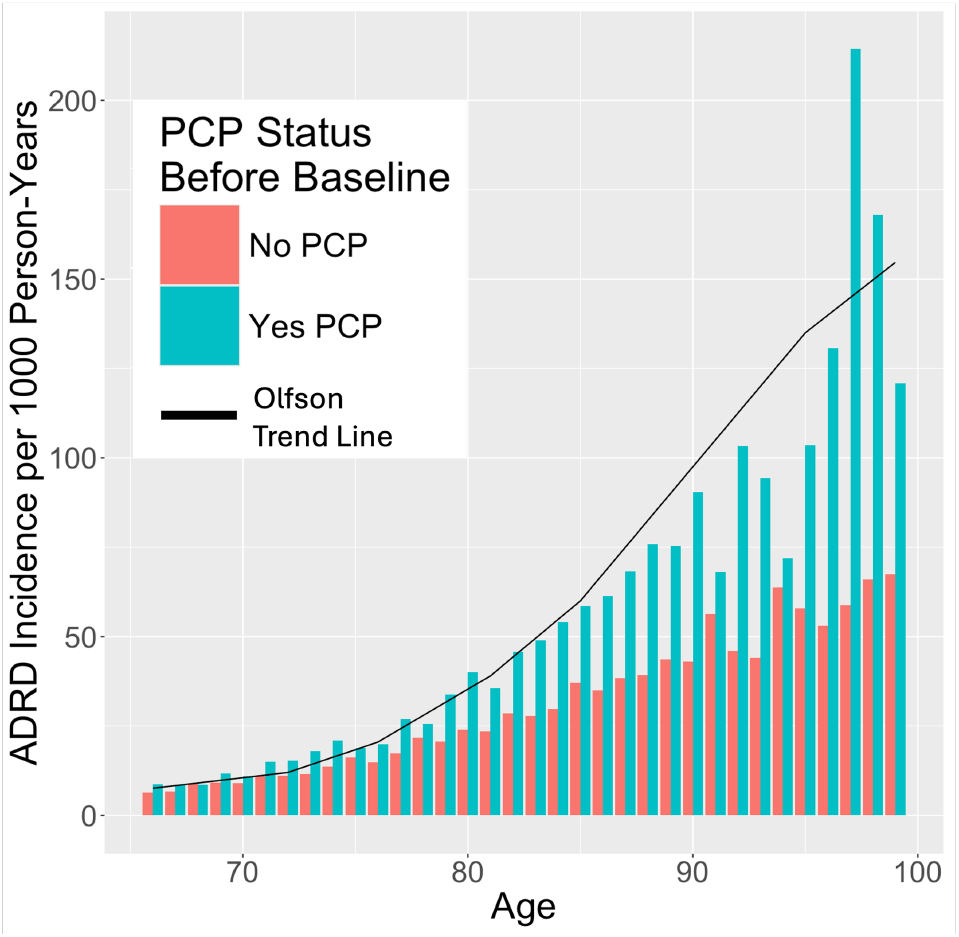
Age-specific ADRD diagnosis incidence per 1,000 person-years comparing PCP vs. no-PCP patients. Black trend line is constructed by interpolating data extracted from a figure reported in a study of over eight-million patients using Medicare claims data (Olfson et al., 2021).

**Figure 5:**
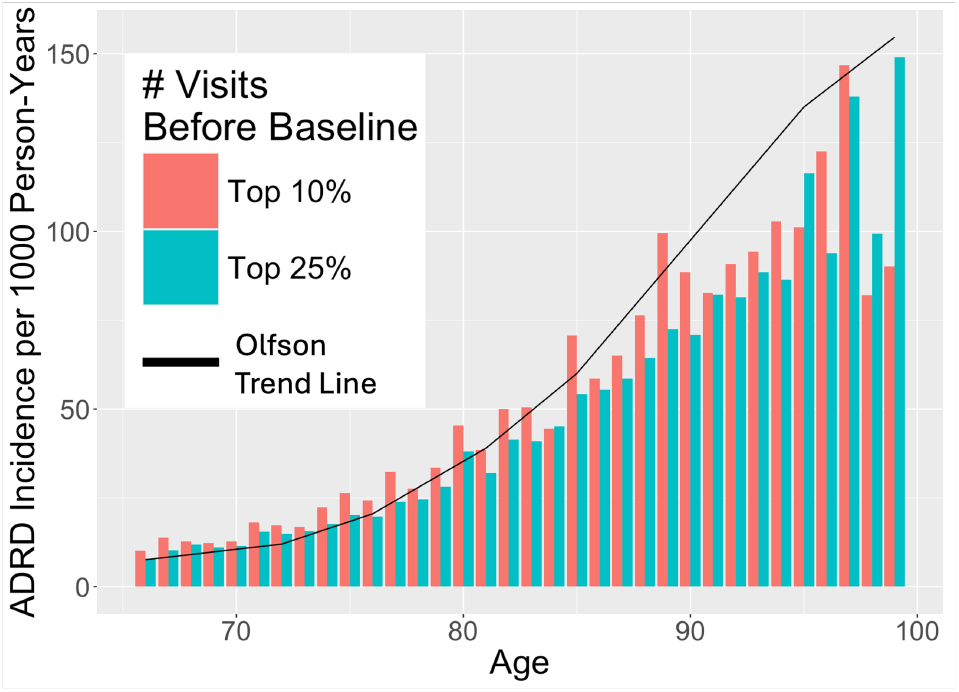
Age-specific ADRD diagnosis incidence per 1,000 person-years comparing patients with the top quartile vs. top decile of visits prior to antidiabetic prescription. Black trend line is constructed by interpolating data extracted from a figure reported in a study of over eight-million patients using Medicare claims data (Olfson et al., 2021).

In an RCT with ADRD as the primary outcome, outcomes are generally observed at regular screenings and ascribed to the date of the screening. Although patients may develop ADRD at any point between screenings, the consistent screening schedule ensures that the delay between the incidence of an outcome and its ascertainment is independent of the treatment arm. Furthermore, by selecting a sufficient screening frequency, the variance caused by the ascertainment delay is minimized.

Because emulating a target trial relies on past healthcare encounters as retrospective screening opportunities, there cannot be a pre-specified schedule common to all patients. Still, by using metrics associated with healthcare utilization in the eligibility criteria and/or propensity score and outcome models, an emulated target trial can induce a group of patients with improved frequency and balance across treatment arms of visits when their medical records are accurately updated. To evaluate the effect of the PCP criterion on the frequency of these informative visits, we first compared the visit frequency of the PCP vs. no-PCP patients, and then examined the value of information added to their EHR at those visits.

For each patient, we quantified the frequency of their visits using the longest continuous time during which the patient did not have a 4-month period without any encounters (*LCT*_4_) (Figure 6). This metric was chosen to reflect a patient’s longest period of uninterrupted care: during this period, ADRD-related outcomes have an opportunity to be recorded within 4 months of symptom manifestation, so ascertainment delays should be similar to those in a clinical trial. We reasoned that 4 months was appropriate because all patients in the cohort have T2D, a chronic disease that generally requires routine follow-up visits every 3 months for prescription renewal. Additionally, sensitivity analyses using other time spans ranging from 1 to 24 months yielded similar results. On average, PCP patients had an *LCT*_4_ of 5.4 years (25%-75% IQR 2.4 - 7.2), while no-PCP patients had an *LCT*_4_ of only 2.4 years (25%-75% IQR .53 - 3.3). Of note, the average timespan between the patient’s first EHR record and their most recent one was greater in the PCP vs. no-PCP group (20.5 yrs vs. 15.9 yrs), but the pronounced difference in *LCT*_4_ held true even after accounting for differences in EHR entry dates by comparing patients whose first record in the EHR is in the same year.

**Figure 6:**
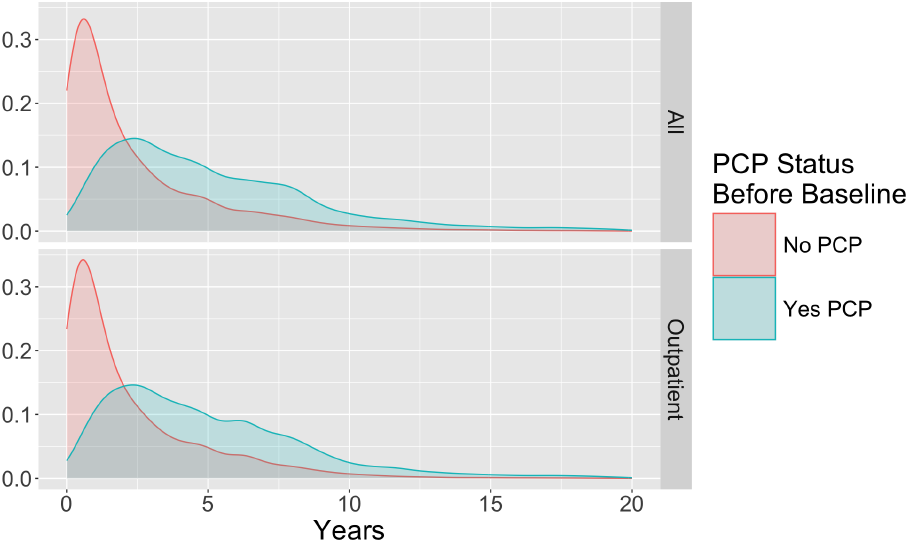
Distribution of each patient’s longest continuous stretch without a 4-month gap between visits (top) or outpatient visits (bottom), stratified by PCP status. The same trend exists for 1, 2, 6, and 12 months.

It is challenging to gauge the chance than an ADRD outcome is assessed and recorded at a given visit without a carefully validated dataset. To address this limitation, we developed a proxy method that relies on disease progression. Patients with mild ADRD who subsequently develop moderate or severe AD generally progress in 3-4 years (Vermunt et al., 2019), so patients with a diagnosis of moderate or severe ADRD (and visits during the previous year) are likely to also have a diagnosis of mild ADRD, as there were opportunities in prior visits for assessments of their cognitive function. Therefore, if a patient in our cohort has a dementia diagnosis and healthcare encounters in the year beforehand, their chance for outcome ascertainment through the visit process can be roughly assessed by whether they have a prior diagnosis of memory loss or mild cognitive impairment (MCI) in their record. Among the 236,025 patients that meet these criteria in our unfiltered cohort, 30.3% of patients in the PCP group have a diagnosis of memory loss or MCI before receiving an ADRD-related diagnosis; in contrast, only 17.8% of patients in the no-PCP group have such a record.

Beyond the stark difference between the PCP and no-PCP groups, the percentage of patients with records of both disease stages remains low overall. Because many patients receive only a single ADRD diagnosis, we use a broad set of codes to define the outcome of ADRD (including both MCI and advanced dementias, which reflect different clinical disease stages). While this composite definition improves the capture of ADRD outcomes, it may affect the estimation of treatment effects if there is a disparity between treatment arms in the stage of ADRD at which the outcome is first recorded. Among patients with ADRD outcomes, far more PCP patients’ outcomes are recorded first with a memory loss or an MCI diagnosis (Figure 7), revealing another disparity that necessitates addressing PCP status.

**Figure 7:**
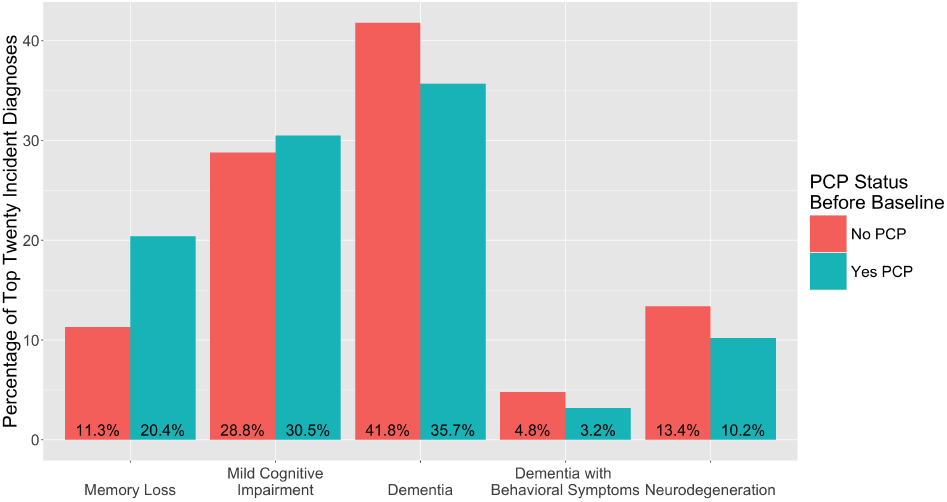
Percentages of the top twenty incident ADRD diagnoses categorized by five classes, grouped by PCP status before baseline. The first two are considered mild, and may suggest earlier recognition of ADRD.

### 2.4. Methods of utilizing the PCP indication

To investigate the sensitivity of TTE results to the approach used to handle the PCP indication, we replicated the prior metformin vs. sulfonylureas emulation. In the original study, 13,191 participants from the MGB EHR with incident antidiabetic prescriptions between January 2007 and September 2018 were selected. From this cohort, patients were excluded if they were younger than 50 at baseline, initiated antidiabetic polytherapy, had no visits in the 18 months before baseline, had an ADRD indication prior to baseline, or had chronic kidney disease (a contraindication for metformin). The covariates used in the propensity model are listed in appendix B.2.

In addition to a baseline strategy ignoring PCP indications (B), we tested two sensitivity analyses comparing methods of using the PCP indication to increase data quality: a modeling strategy adding a PCP covariate in the propensity model (M), and an exclusion strategy requiring a PCP indication for inclusion in the cohort (E). To compare these strategies to another proposed healthcare utilization metric (Goldstein et al., 2016), we also tested two strategies where the number of visits a patient had prior to baseline was added as a feature in the propensity score and outcome models. In the Visits (V) strategy, there was no usage of the PCP inclusion criterion, and in the Visits-Exclusion (V-E) strategy, the visits modeling feature was used with the PCP inclusion criterion. Aside from the use of these indications, our study diverged from the original methodology by including data through April 2024. (B), (M), and (V) had a cohort of N=54,638 (46,714 metformin, 7,924 sulfonylureas) and (E) and (V-E) had N=17,112 (15,904 metformin, 1,208 sulfonylureas). Although we didn’t perform a power analysis, all of our cohorts were larger than that of the original study.

The PCP strategy was evaluated across these five methodologies by comparing the hazard ratio (HR) obtained from a Cox proportional hazards model, and cumulative incidence functions (CIF) that account for the competing risk of death (Getzen et al., 2023).

## 3. Results

We found that the HR estimates for the effect of initiating metformin were similar in (B) and (M): .87 (95% CI: .78 - .96, p=.006) and .85 (95% CI: .77 - .95, p=.003), respectively (Figure 8). In (E), the HR estimate was .76 (95% CI: .61 - .95, p=.016), with an expected increase in confidence interval width, given the much smaller cohort size after selecting on the PCP indication. However, across 5,000 random samplings of 15,910 metformin initiators and 1,208 sulfonylureas initiators from (B), only 19 had a CI width as small as (E). Therefore, there is less of an increase in confidence interval width in (E) than expected, suggesting that the exclusion of no-PCP patients may reduce some noise.

**Figure 8:**
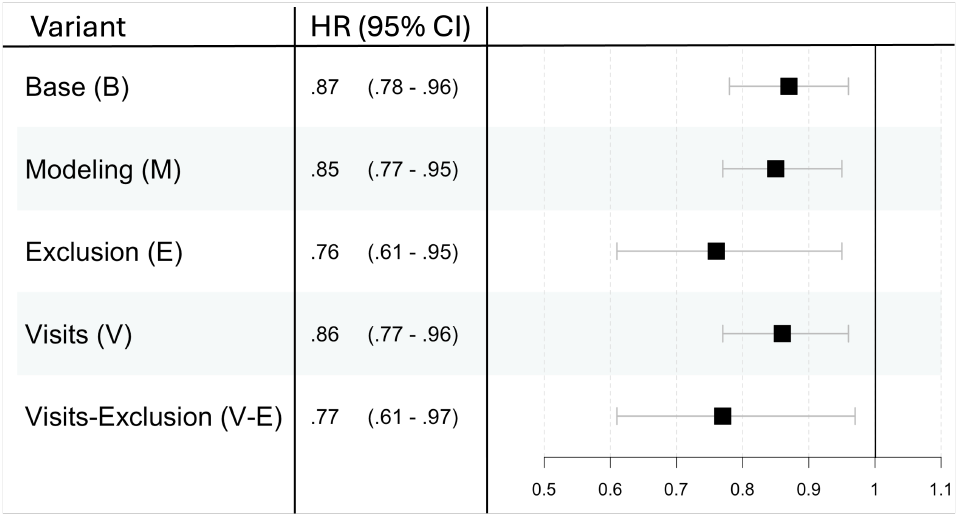
Hazard ratios and 95% confidence intervals for the estimated effect of initiating metformin vs. sulfonylureas on ADRD incidence, estimated from Cox models.

Although the goal of the PCP feature is to improve the model’s reflection of a real-world effect and not specifically to increase model performance, we note that the concordance indices are highest in (E) for both the ADRD (.73 vs. .72 in B and M) and ACM (.78 vs. .76 in B and M) models. The stronger effect in (E), which is outside the confidence intervals of every experiment without the PCP exclusion criterion, may be due to reducing confounding by eliminating patients with less complete and accurate EHR data. That being said, the confidence intervals across all 5 experiments do overlap with each other and are always below 1, robustly supporting the positive effect of metformin vs. sulfonylurea treatment initiation.

For both (V) and (V-E), the visits feature appears to have a minimal impact. The similarity between (B), (V) and (M) – as well as between (E) and (V-E) – suggest that the use of an exclusion criterion influences the estimated treatment effect more than one additional propensity score and outcome model feature.

To quantify the treatment effect accounting for the competing risk of death, the risk difference across treatment arms at ten years after baseline (*RD*_10_) was examined. Plots for the risk difference from 0-15 years for both death-without-dementia (DØD) and ADRD can be found in Appendix E. With the addition of the PCP status covariate to the propensity model, the DØD curves for the two arms are slightly closer in (M) than (B): the *RD*_10_ of DØD were -8.0% (95% CI: -9.1% - -7.2%) and -9.7% (95% CI: -10.1% - -8.8%), respectively (Figure 9). Having an internal PCP is more common among metformin patients, so this suggests that decreasing the weight of metformin patients with a PCP indication mildly attenuates the mortality survival-time effect. For the ADRD curves, the *RD*_10_ are -.29% (95% CI: -.98% - .56%) in (B) and -.52% (95% CI: -1.3% - .34%) in (M), so the elimination of residual confounding from disproportionality in PCP status reveals a slightly more pronounced effect of metformin (Figure 9).

**Figure 9:**
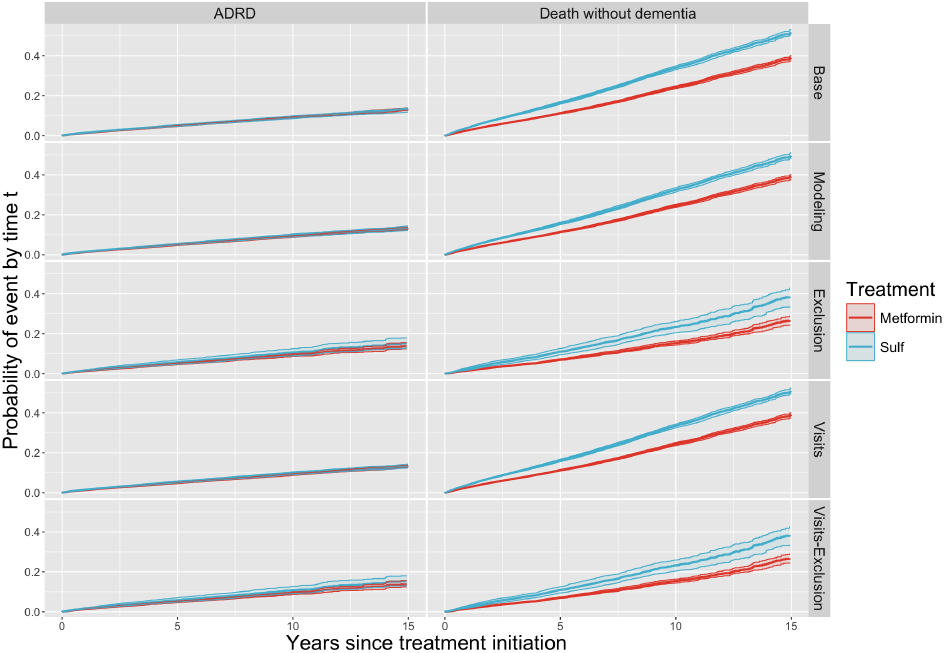
Cumulative incidence of ADRD (left) and death-without-dementia (right) comparing the sulfonylureas (blue) and metformin (red) arms for each strategy.

In (E), the ADRD curves for both treatments increase (Figure 9), which is far more likely to be the result of higher outcome ascertainment in the entire cohort than of a higher rate of actual cognitive decline among patients with a PCP indication. As observed in the Cox model, the smaller sample size results in much larger confidence intervals. In this experiment, the *RD*_10_ of DØD is -8.0% (95% CI: -10.8% - -4.3%), a similar result as in (M). Notably, the *RD*_10_ of ADRD is -1.4% (95% CI: -3.0% - .22%), more than four times that found in (B).

Interestingly, the visits feature had a more noticeable effect on the risk differences than on the hazard ratios. In (V), the *RD*_10_ of DØD was -9.2% (95% CI: -10.2% - -8.5%) and the *RD*_10_ of ADRD was -.65% (95% CI: -.15% - .01%), revealing that the inclusion of the visits feature in the propensity score model was more impactful on the ADRD risk – but less impactful on DØD risk – than the use of the PCP feature in the same way. In (V-E), the *RD*_10_ of DØD was -7.8% (95% CI: -10.6% - -4.2%), a similar result to (E). The *RD*_10_ of ADRD was -1.5% (95% CI: -3.2% - .13%), the strongest risk difference found in any strategy.

Although the most influential factor overall was the use of the PCP status in the exclusion criteria, using the visits feature in modeling showcases the possibility of using multiple strategies to account for healthcare utilization. PCP status – being a binary variable – cannot entirely capture disparities between patients’ data quality, and the visits feature fails to account for different types of encounters and their distinct effects on the EHR record. Using both features can establish a better balance in covariate measurement and outcome ascertainment between the two treatment arms, and the addition of further healthcare utilization metrics may continue to alleviate residual confounding.

As with the hazard ratios, the confidence intervals of the *RD*_10_ produced in each strategy overlap with each other. This supports the positive impact of metformin initiation, and suggests that while the use of healthcare utilization metrics is critical for producing accurate point estimates, it is unlikely to alter the direction of an effect.

## 4. Discussion

### 4.1. Practical Implications

We found that the patients with an internal PCP appeared to have far less missing data across death records, diagnosis records, and lab results. This population also aligned well with the general population in age-specific mortality and ADRD incidence rates, unlike the population of patients with records in the EHR but no internal PCP. Given the improved data quality and alignment with the target population, the PCP feature is an important consideration when executing an EHR-based TTE, and its use significantly impacted the results of the metformin vs. sulfonylurea trial, especially when used as an eligibility criterion.

The best use for the PCP feature depends on the particular trial. Including internal PCP status in the propensity model can eliminate confounding effects caused by an imbalance in PCP status across treatment arms. However, all healthcare utilization metrics are potentially affected by treatments and outcomes, and can often act as a collider (Weiskopf et al., 2023). Fortunately, the PCP feature should result comparatively minimal bias, as primary care encounters are less correlated to specific medical conditions or events (Weiskopf et al., 2023). Also, if a large portion of the population does not have an internal PCP, then retaining them in the cohort weakens the assumption that the EHR records correspond well to actual events. This reduction in confidence is deceptive because it will not materialize in the results as lower significance, which measures confidence in the signal between records regardless of their correlation to actual events.

Using the PCP feature as an eligibility criterion addresses this issue by removing the lower quality data. However, as with any eligibility criterion, it reduces the power of the trial by diminishing the cohort size. This filtering also potentially introduces selection bias because the internal PCP population may be skewed across various features; for instance, patients in the US with a PCP have higher educational attainment and socioeconomic status overall (Getzen et al., 2023).

In our metformin vs. sulfonylurea TTE, every patient is receiving an antidiabetic prescription in the US and is therefore almost certain to have a PCP somewhere. In this case, the primary distinction of the PCP cohort is that their PCP is internal to the EHR, minimizing the potential selection bias from removing patients without a PCP anywhere, which is corroborated by the alignment between our PCP cohort and the general population. Additionally, the clinics in our EHR network have many patients that come only for tertiary care, resulting in many patients that only have data related to those specific visits. The EHR also has thirteen million patients, providing a large starting cohort. Therefore, we believe that the exclusion strategy is most useful for this emulation. For other TTEs, the best use of the PCP criterion requires a thoughtful consideration of the full eligible population and the clinics in the EHR network.

### 4.2. Related Work

There has been a lot of work characterizing many data quality issues. The challenge is especially prevalent in the context of a TTE with ADRD as the primary outcome, given that most dementia phenotyping algorithms have a PPV or NPV below 70% (Walling et al., 2023). Prior work has also reviewed missingness in key covariates among diabetic populations; for instance, a review from February 2025 of EHR data from Spain found that 19.9% of diabetic patients had missing blood pressure measurements and 35.4% had missing BMI records, both of which are important features in a propensity model (Quesada and Orozco-Beltran, 2025). The problem of data missingness – and the detrimental impact on TTE and observational studies using EHR data – is well established, and there has been an effort in the community to ensure that studies using the EHR address data quality directly (Haneuse et al., 2021).

Given the strong potential for using EHR data, many methods have been proposed for handling data quality concerns. When possible, external data can be used to supplement healthcare records Wells et al. (2013), a strategy we use with MA death records. The bias caused by missing data can be minimized through careful patient censoring Wells et al. (2013); for example, a patient’s last encounter date should be determined by their final completed visit to ensure that canceled visits or phone calls are not treated as follow-ups. For ADRD outcomes among others, there has been work to extract data from the unstructured physician notes using large language models, with tools such as NAT (Noori et al., 2022).

As discussed, our PCP criterion is not the only utilization-based method to improve data quality, and it should preferably be used in conjunction with other methods. Our visits feature was adapted from Goldstein et al. (2016), who used simulated and EHR data to demonstrate that outcome adjustment on a patient’s number of encounters in the EHR meaningfully changes the odds ratio between the recordings of depression and weight loss. In the novel context of a TTE with ADRD as the primary outcome, the visits prior to baseline noticeably impacted risk differences when accounting for the competing risk of death, although it did not significantly affect the hazard ratio in (V) and (V-E). In future work, we plan to investigate other harmonious utilization metrics.

### 4.3. Limitations

In this study, we treated the PCP feature as a binary label. This simple representation does compress more detailed information about the patient’s primary care relationship; for instance, patients can have varying levels of loyalty to their PCP, or have a specialist that they see regularly who acts in some ways like their PCP. In the future we would like to explore these intricacies.

Despite the potential variation between types of PCP relationships, having a binary representation allows the PCP criterion to naturally act as an inclusion criterion. While we did test features based on a patient’s number of various types of visits, there was no clear motivation to select a specific cutoff for excluding patients. In our tests, we investigated cutoffs at the quartiles in order to get a general sense of the possible effects of each alternative healthcare utilization criterion, but in any given TTE, it would be difficult to rationalize a particular choice. Our TTE results were highly sensitive to the cutoff: selecting patients in the top three quartiles of visits prior to baseline resulted in a hazard ratio of .87, but selecting only those in the top quartile resulted in a hazard ratio of .77. Filtering on the number of visits prior to baseline does improve data quality, but it necessitates pre-specifying a reasoned cutoff parameter, unlike the PCP criterion.

Because this study used a single EHR and cohort, it is important to investigate the generalizability of the results in order to more broadly evaluate the utility of the PCP feature. With different populations, the effect of PCP status may differ, so we are replicating this work in a hypertensive cohort from the same EHR. Additionally, we are exploring the use of a similar feature in a geographically distant EHR, where differences in demographics, clinical practices, and EHR platforms may impact both the implementation and benefits of a PCP feature.

Countries with different healthcare systems may not have analogs to an internal PCP, the PCP criterion can still benefit collaborations between groups working with EHRs in these countries and groups working with EHRs in the USA. Countries with a national healthcare system often have a closed EHR network and patients with more regular healthcare encounters, so selecting a cohort of patients in the USA with the internal PCP eligibility criterion may provide similar qualities and improve harmonization between sites.

While EHR data are a valuable source of information, EHR studies are inherently unrepresentative of the general population. A key step in working towards generalizability is reporting demographic data more comprehensively (Boyd et al., 2023), which can be more effective when using the PCP feature. Because EHR data are disproportionately missing for less privileged groups, the demographics of patients with *any* records in the EHR differs from the demographics of patients with *thorough* records, so ignoring the PCP criterion may obscure the latent bias. Future work to improve data quality should further refine the PCP criterion, such as by incorporating unstructured provider notes in the EHR. To mitigate biases in the EHR and fully benefit from the PCP criterion, studies should expand generalizability through replications in other EHRs, and more broadly with public efforts to promote healthcare accessibility for marginalized groups.

### 4.4. Future Steps

To formulate our definition of internal PCP encounter, we chose indications based on a manual review of the codes present in RPDR. In the future, we would like to refine our PCP label assignment by using a dataset of validated PCP labels to tune our use of the available indications. We also will to investigate the unstructured physician notes, which may be identify more patients with internal PCPs, and thereby reduce the loss in power caused by the cohort reduction.

Other information within the physician notes may decrease data missingness and improve accuracy for all patients, and possibly change the effect of using the PCP criterion. We hypothesize that patients with an internal PCP would benefit most from the additional data because their PCP’s notes specifically are likely to contain thorough medical histories. We plan to evaluate this idea by introducing data both through chart reviews performed by expert clinicians and potentially using large language models to phenotype text from unstructured physician notes.

## Data Availability

The study uses EHR data from the Research Patient Data Registry (Nalichowski et al., 2007), so- cial vulnerability index (SVI) data from the Agency for Toxic Substances and Disease Registry (https://www.atsdr.cdc.gov/placeandhealth/svi), and Massachusetts death records from the Registry of Vital Records and Statistics. Because the data contain patient information, they cannot be made available.

## 5. Citations and Bibliography

## Acknowledgments

The authors thank other members of the DRIAD-EHR team including Bella Vakulenko-Lagun, Deborah Blacker, Sudeshna Das, Jeff Klann and Shawn Murphy for thoughtful comments. This study was funded by the NIH R01 AG058063 (awarded to M.W.A.) and by supported by the Eric and Wendy Schmidt Center at the Broad Institute of MIT and Harvard.

