## Appendix A - H for "Investigating Primary Care Indications to Improve the Quality of Electronic Health Record Data in Target Trial Emulation for Dementia"

### Appendix A. Table by Treatment Arm

Summary statistics, comparing patients who initiated on metformin vs. sulfonylureas. Covariates are measured at baseline, the date of the patient's first prescription of metformin or a sulfonylurea. Outcomes of ADRD and death are prevalences after baseline. For age, social vulnerability scores (SVS), and visit metrics, means are reported with standard deviations. For sex, education (ed) levels, and covariate diagnoses (Dx), percentage values are reported.

Table 1

| **Feature** | **Metformin** | **Sulfonylurea** |
|---|---|---|
| Patients | 46,714 | 7,924 |
| PCP | 15.2% | 34.0% |
| ADRD | 4.9% | 8.8% |
| Death | 14.2% | 37.3% |
| Age at Baseline | 65.1% (9.0) | 70.8% (10.8) |
| Sex Female | 51.6% | 47.1% |
| Ed. Secondary | 32.9% | 35.9% |
| Ed. College | 37.5% | 30.1% |
| Ed. Graduate | 8.3% | 5.3% |
| Ed. Missing | 21.4% | 28.7% |
| Socioeconomic SVS | 0.33 (.22) | 0.33 (.21) |
| Home Life SVS | 0.450 (.20) | 0.46 (.20) |
| Racial/Ethnic SVS | 0.44 (.22) | 0.42 (.22) |
| Housing SVS | 0.51 (.18) | 0.51 (.18) |
| Hypertension Dx | 62.9% | 65.8% |
| Stroke Dx | 4.6% | 6.4% |
| Cancer Dx | 32.3% | 34.0% |
| COPD Dx | 3.7% | 4.1% |
| Overweight Dx | 10.4% | 4.6% |
| Obesity Dx | 33.6% | 20.3% |
| CVD Dx | 18.3% | 27.9% |

Table 1

| Feature | Metformin | Sulfonylurea |
|---|---|---|
| HbA1c Missing | 41.0% | 48.4% |
| HbA1c Reference | 4.7% | 4.1% |
| HbA1c Prediabetes | 18.3% | 11.1% |
| HbA1c Diabetes | 36.0% | 36.4% |
| BP Missing | 62.6% | 82.5% |
| BP Reference | 17.1% | 7.6% |
| BP Hypertension 1 | 9.5% | 4.2% |
| BP Hypertension 2 | 10.8% | 5.8% |
| Total Visits | 66.2 (94.3) | 46.8 (71.8) |
| Outpatient Visits | 58.6 (87.6) | 37.1 (63.3) |
| Years in EHR | 17.6 (7.8) | 15.7 (7.7) |

### Appendix B. TTE Details

#### B.1. Outcome Definitions

ADRD outcomes were defined as the first occurrence of any of the following ICD9, ICD10, internal diagnosis codes, or medications indicating cognitive decline. These sets were developed by consultation with expert clinicians.

The ICD9 codes used were the following:
294.8, 290.40, 294.20, 294.21, 290.0, 294.10, 331.83, 331.9, 294.0, 294.9, 290.13, 331.3, 331.0, 331.5, 331.2, 331.82, 290.43, 290.21, 290.10, 780.93, 290, 331, 294, 294.1, 290.41, 290.3, 294.11, 290.20, 290.42, 290.4, 291.2, 290.11, 331.11, 331.89, 290.9, 331.1, 331.19, 331.7, 290.12, 290.0.1, 290.21.1, 290.20.1, 290.40.1, 290.1, 294.80.1, 294.10.1, 290.10.1, 292.82, 290.3.1, 331.2.3, 331.0.3, 290.42.1, 331.81, 290.8, 294.9.1, 290.43.1, 290.2

The ICD10 codes used were the following:
F03.90, F03.91, F01.50, G31.84, F01.C0, G30.9, G30.1, F02.80, G31.83, G31.89, F01.51, F02.B0, F02.81, F01.518, G30.0, F03.918, F03.A0, G31.9, F10.27, G30.8, I69.311, F02.818, G31.09, F01.A0, F02.A0, F03.911, F03.C0, F01.B0, F02.C11, G31.2, F03.B0, F03.B18, F10.97, I69.911, F03.B11, G31.85, F03.92, F02.C0, I69.811, F02.811, F03.C11, F03.A4, F01.A18, G31.01, G31.81, F01.B11, G31.1, F02.C18, F01.52, F03.9, F03.C18, F02.B2, F01.B4, I69.211

The other codes used were the following:
LPA99, YHAL6, WLAG8, WHMT3, LPA1009, WLEN6, LPA1404, LPA730, LPA867

The medications used were the following:
Galantamine (Razadyne, Razadyne ER) Donepezil (Aricept) Rivastigmine (Exelon) Memantine (Namenda)

Death outcomes were determined by death records within the EHR, supplemented with data from the MA death registry.

#### B.2. Covariate List

The covariates used in the propensity model and in the Cox PH model were the following:

Age at baseline

Sex

Hypertension prior to baseline

Stroke prior to baseline

Chronic Obstructive Pulmonary Disease prior to baseline

Overweight diagnosis prior to baseline

Obesity diagnosis prior to baseline

Cardiovascular disease prior to baseline

Cancer (defined with a strict set of ICD codes) prior to baseline

Cancer (defined with a broad set of ICD codes) prior to baseline

Educational attainment level (pre-college, college, graduate, or missing)

Socio-economic vulnerability score (accounts for income, employment, debt, education, etc.)

Home life vulnerability score (accounts for age of family members, size of family, language proficiency in family, other family vulnerabilities, etc.)

Racial/Ethnic vulnerability score (accounts for racial and ethnic minority status)

Housing vulnerability score (accounts for home type, vehicle access, etc.)

BMI classification ($0- < 25$, $25- < 30$, $30+$)

These covariates were used as main effects.

The social vulnerability index score values were determined by the mean SVI values from all the census tracts within the patient's zip code, using data from the Agency for Toxic Substances and Disease Registry (https://www.atsdr.cdc.gov/placeandhealth/svi). Scores range from 0 to 1, where higher scores indicate more vulnerability.

### Appendix C. Full Forest Plots from the Cox PH Models

Forest plots depicting the estimated hazard ratios from the Cox proportional hazards models for all three variants. The first line shows the estimated treatment effect; following lines show estimated hazard ratios for the covariates in the model. We note that the confidence intervals on coefficients do not account for variance in the propensity model weights.

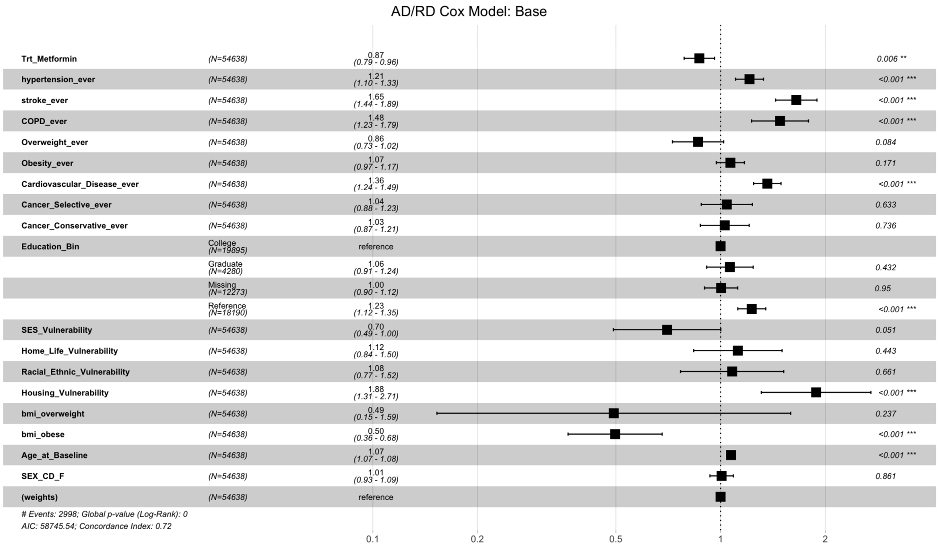


Figure 1: Forest Plot of Cox PH Model (B)

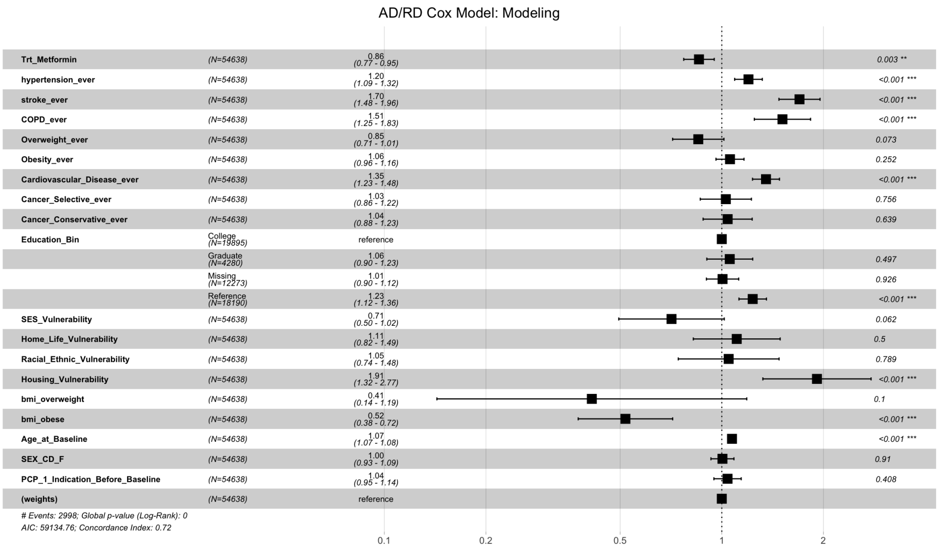


Figure 2: Forest Plot of Cox PH Model (M)

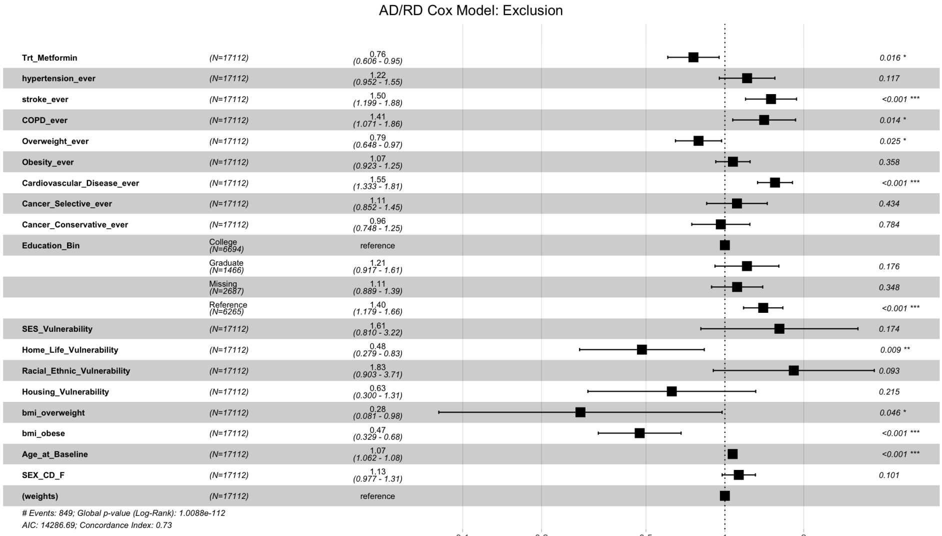


Figure 3: Forest Plot of Cox PH Model (E)

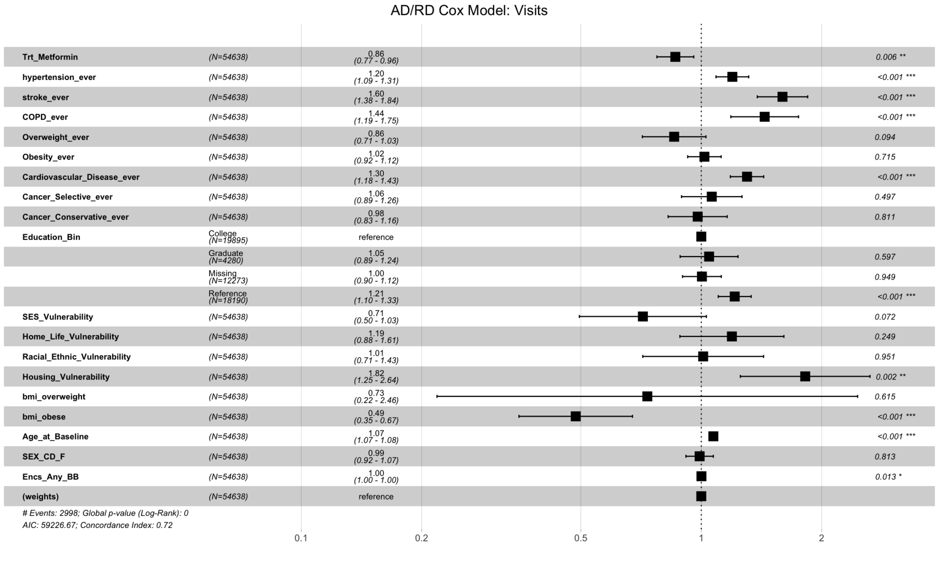


Figure 4: Forest Plot of Cox PH Model (V)

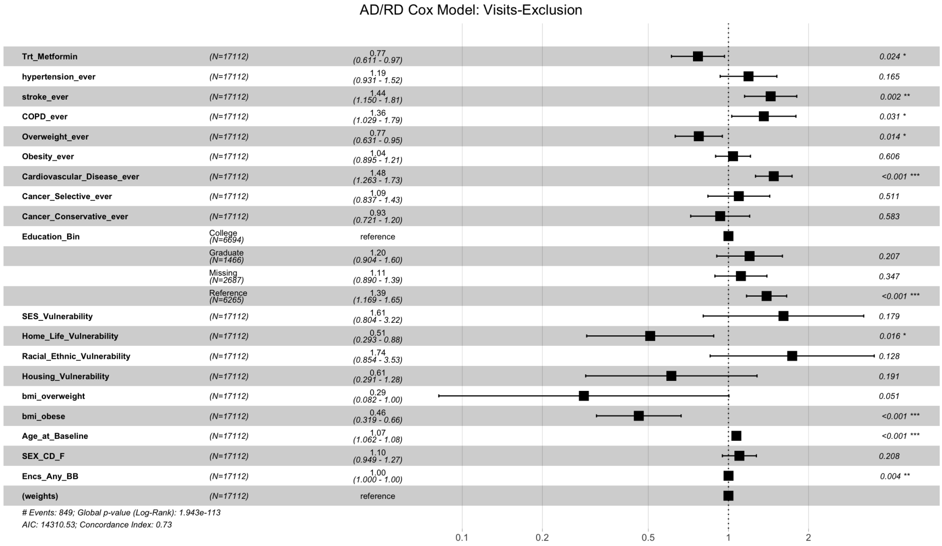

Figure 5: Forest Plot of Cox PH Model (V-E)

### Appendix D. Full-size Individual CIFs

Cumulative incidence functions for both ADRD incidence and death-without-dementia. Plots were produced with the R package CausalCmpRsk (Vakulenko-Lagun et al., 2023), which handles propensity weight variability. Covariates are used to train the propensity score model, but are not used in outcome prediction.

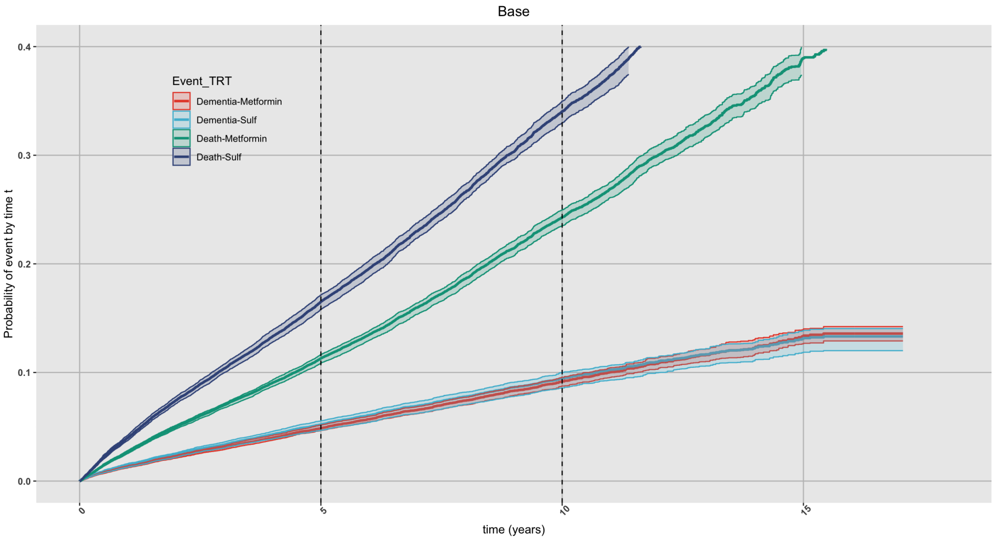


Figure 6: CIFs accounting for competing risks (B)

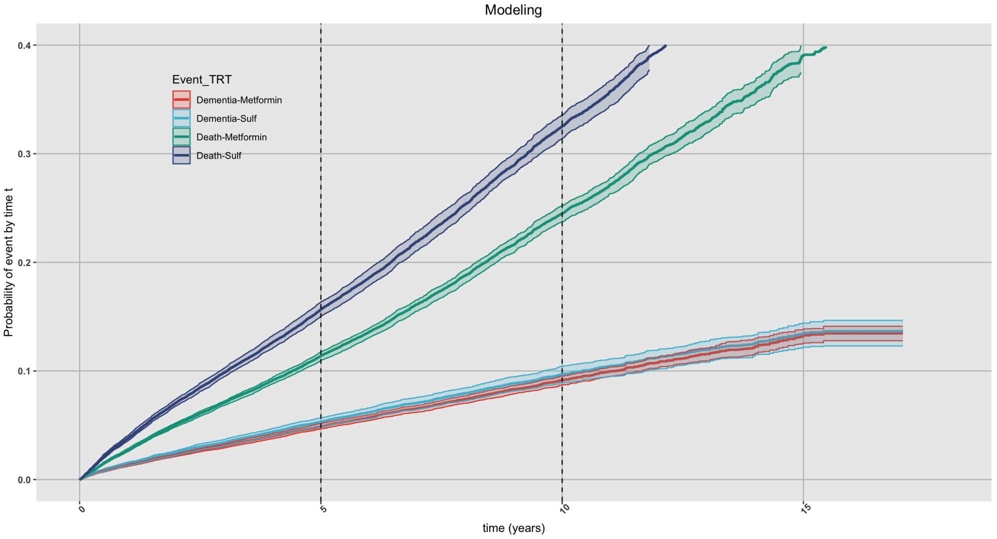


Figure 7: CIFs accounting for competing risks (M)

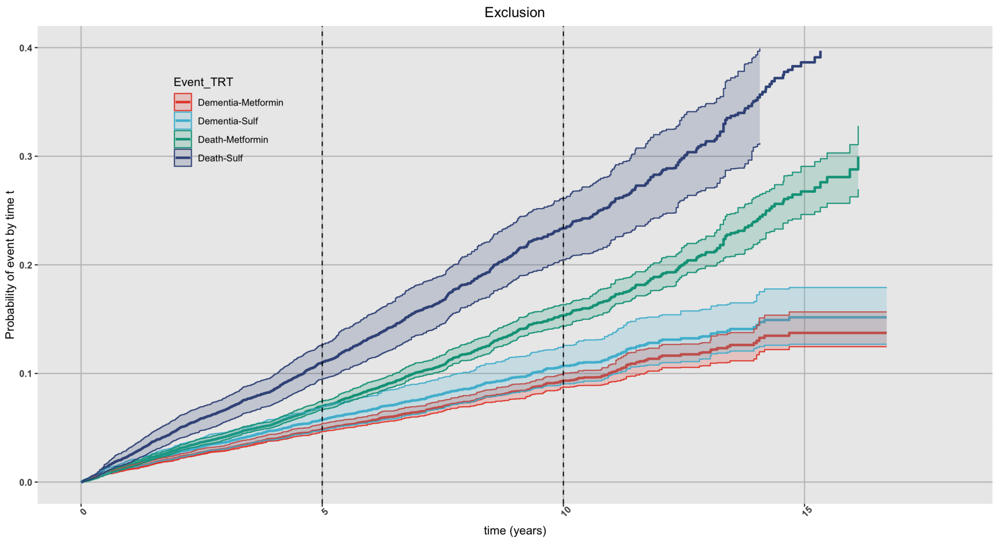


Figure 8: CIFs accounting for competing risks (E)

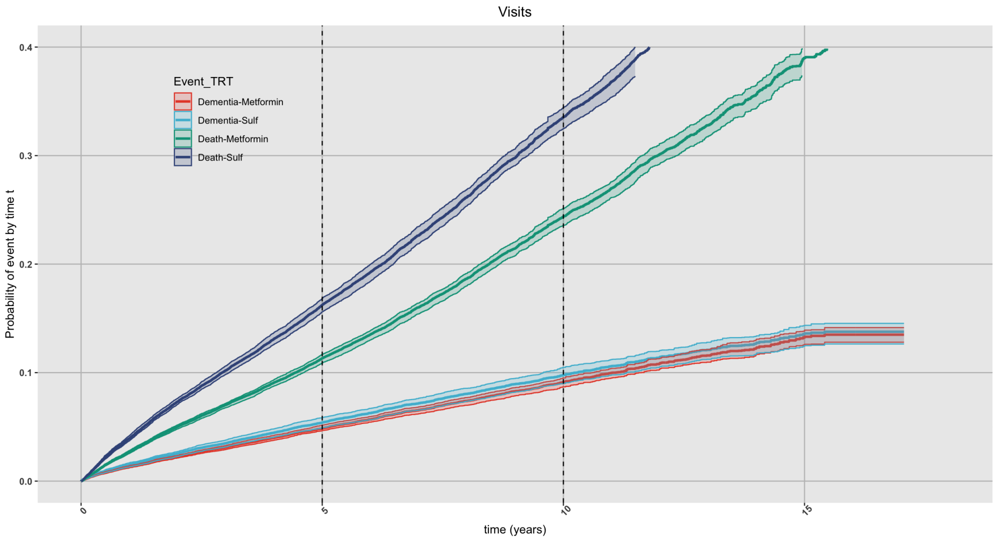


Figure 9: CIFs accounting for competing risks (V)

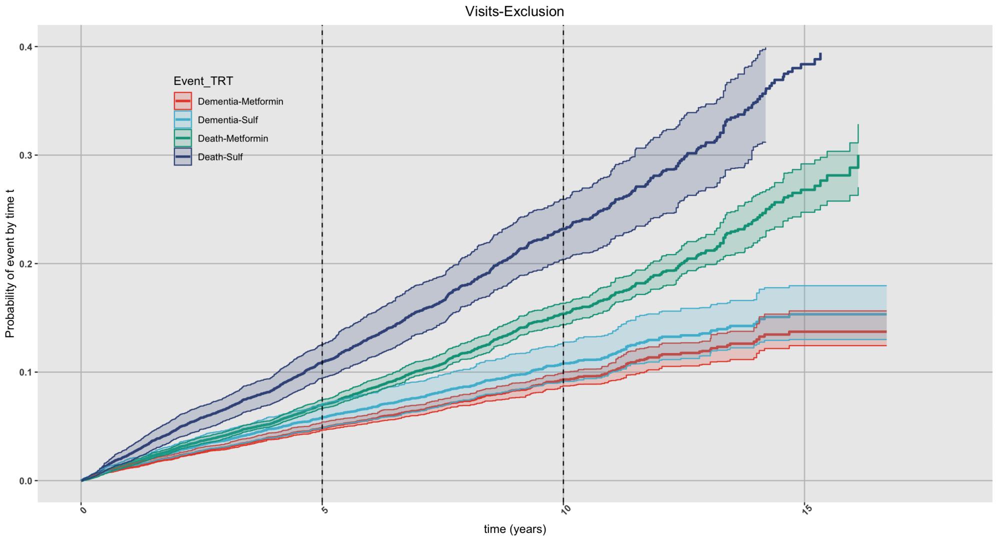


Figure 10: CIFs accounting for competing risks (V-E)

### Appendix E. Risk Difference Plots

Risk differences across metformin vs. sulfonylurea initiators for both ADRD incidence and death-without-dementia. Plots were produced with the R package CausalCmpRsk (Vakulenko-Lagun et al., 2023), which handles propensity weight variability. Covariates are used to train the propensity score model, but are not used in outcome prediction.

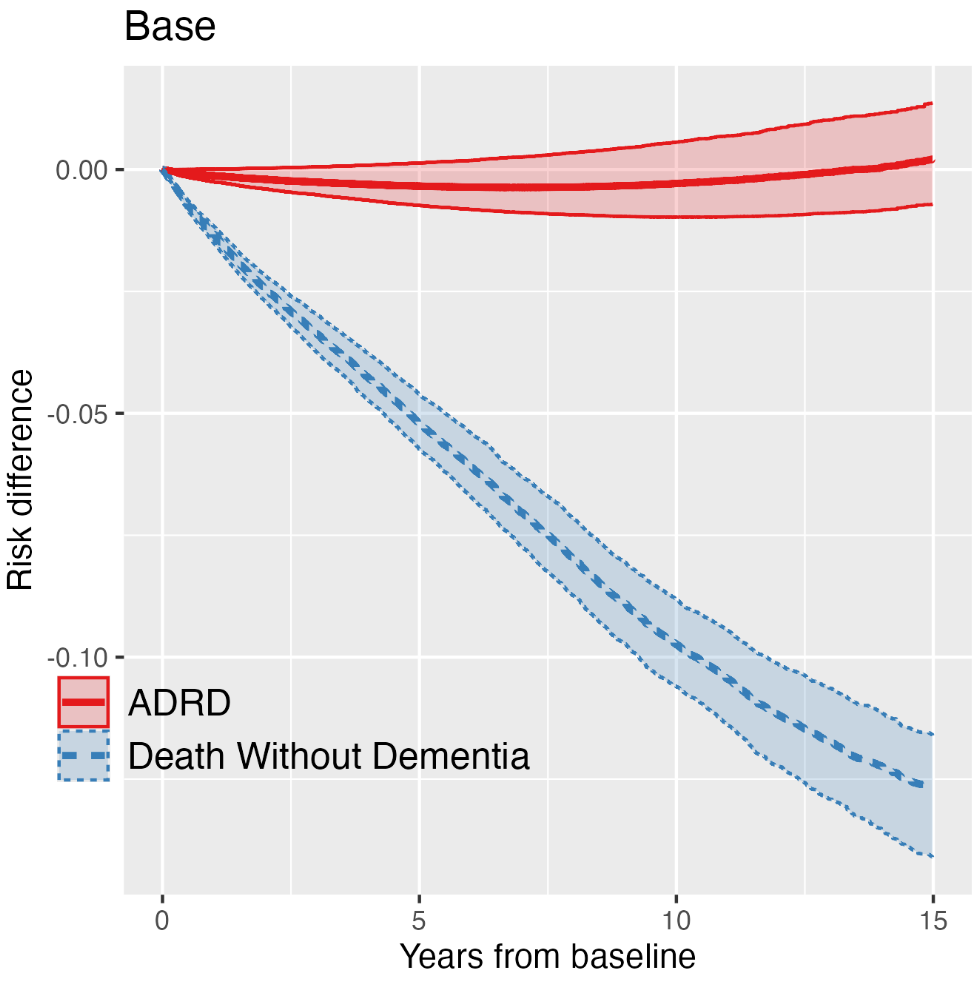


Figure 11: Risk Differences over time for ADRD and DØD (B)

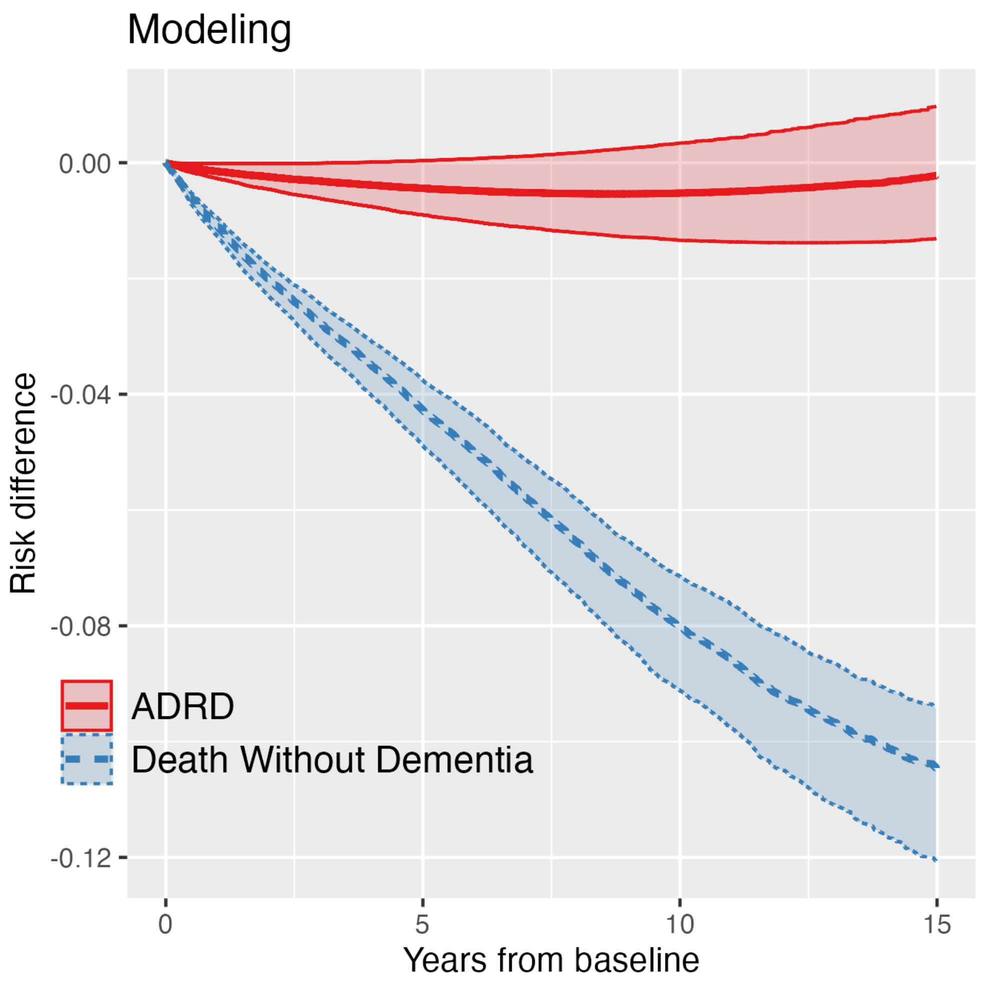


Figure 12: Risk Differences over time for ADRD and DØD (M)

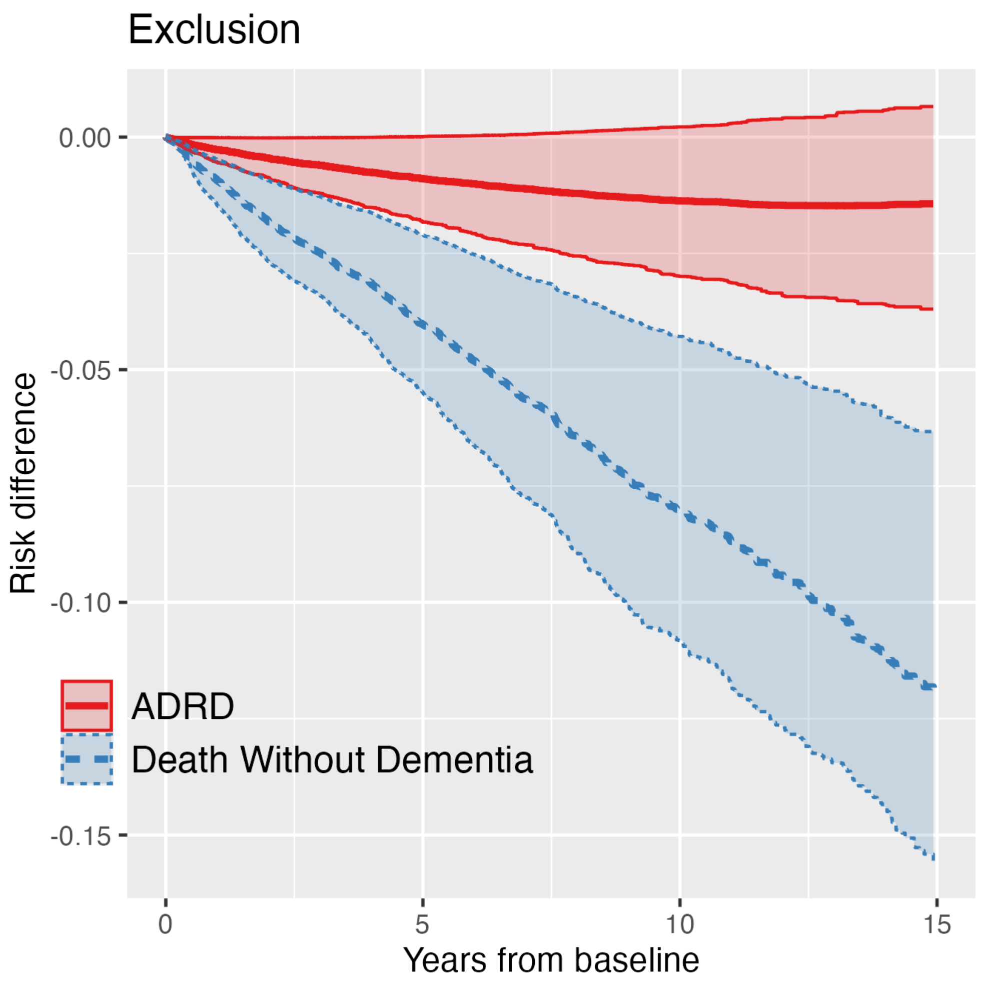


Figure 13: Risk Differences over time for ADRD and DØD (E)

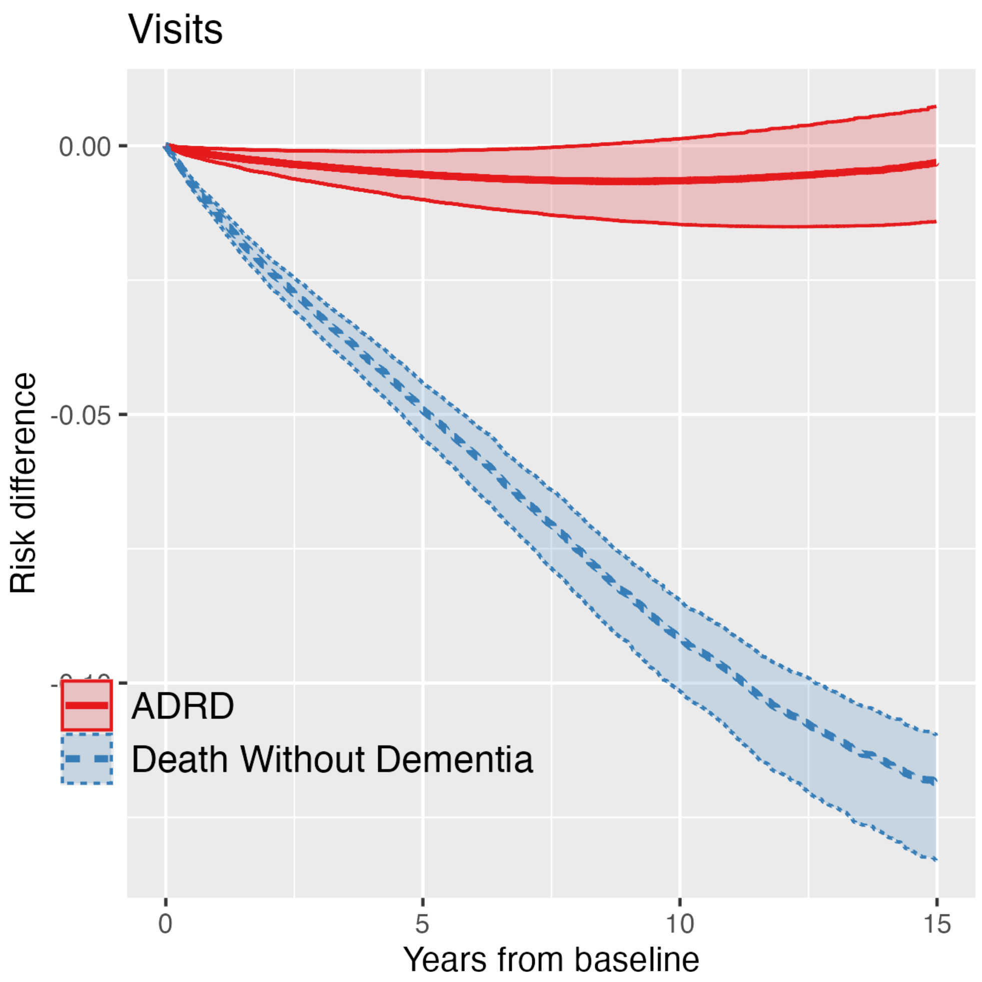


Figure 14: Risk Differences over time for ADRD and DØD (V)

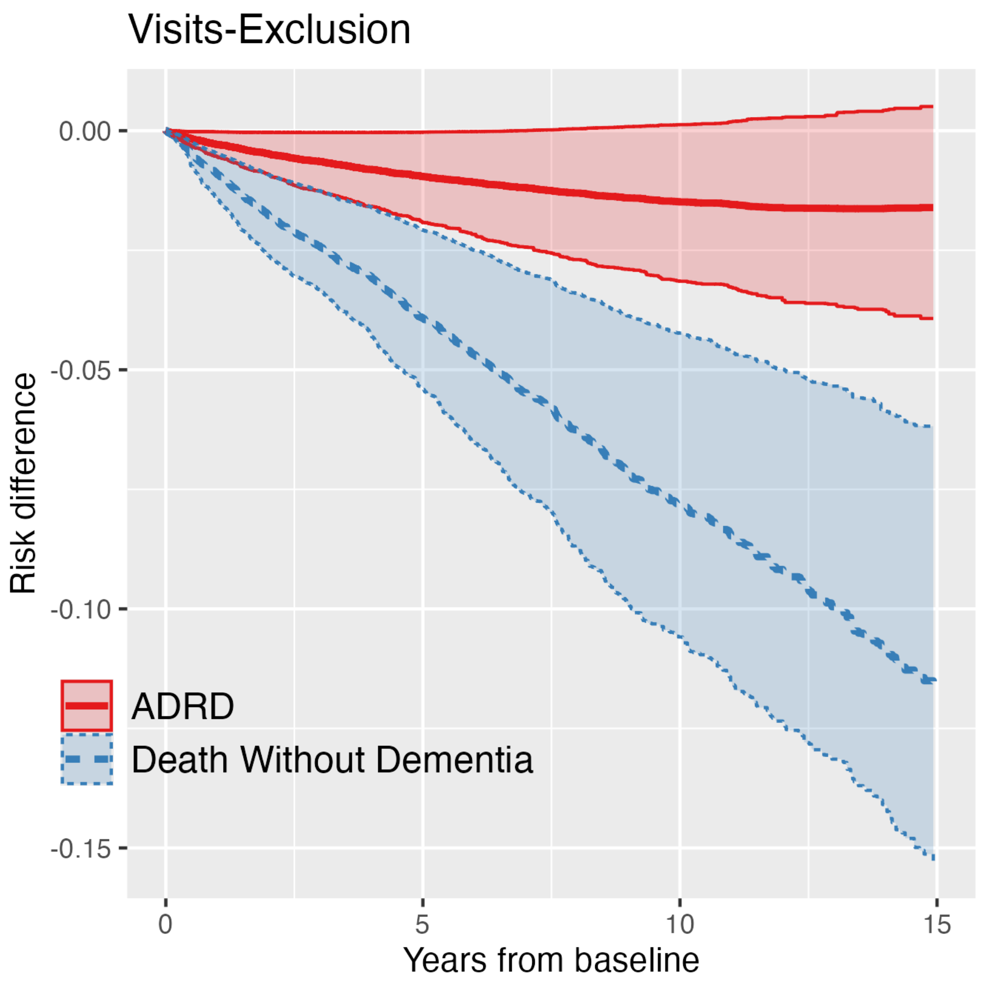


Figure 15: Risk Differences over time for ADRD and DØD (V-E)

### Appendix F. Propensity Score Distributions

Plots showing the distribution of propensity scores grouped by treatment arm. When a score is closer to 1, the model is more confident that the patient is in the metformin arm. The distribution for metformin patients (red) is closer to the right than that of the sulfonylurea patients (blue), but there is significant overlap, suggesting that the two arms are composed of comparable populations. To calculate weights from the propensity scores, we use stabilized average treatment effect weighting (as did the original metformin vs. sulfonylureas TTE).

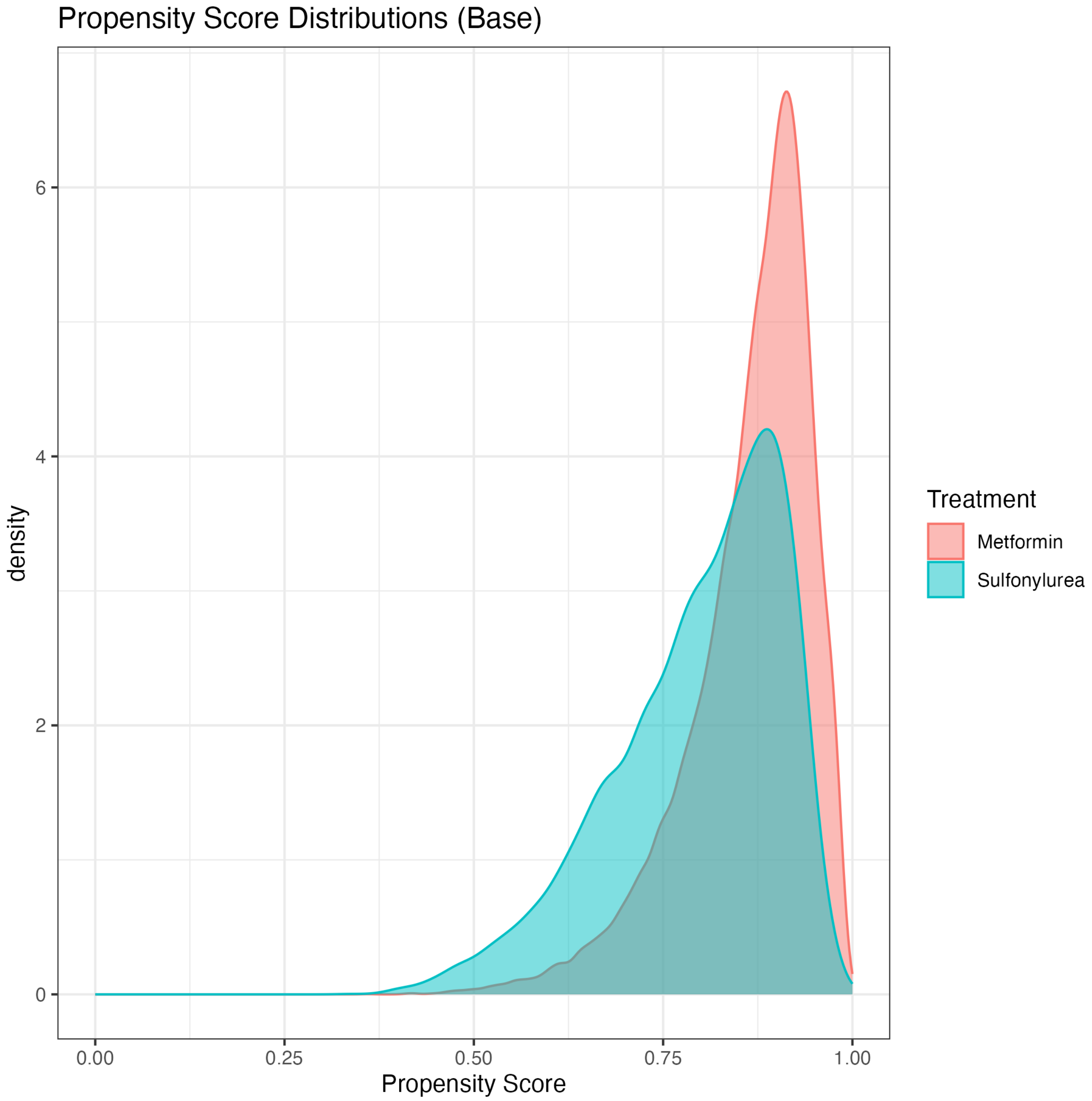


Figure 16: Propensity Distributions (B)

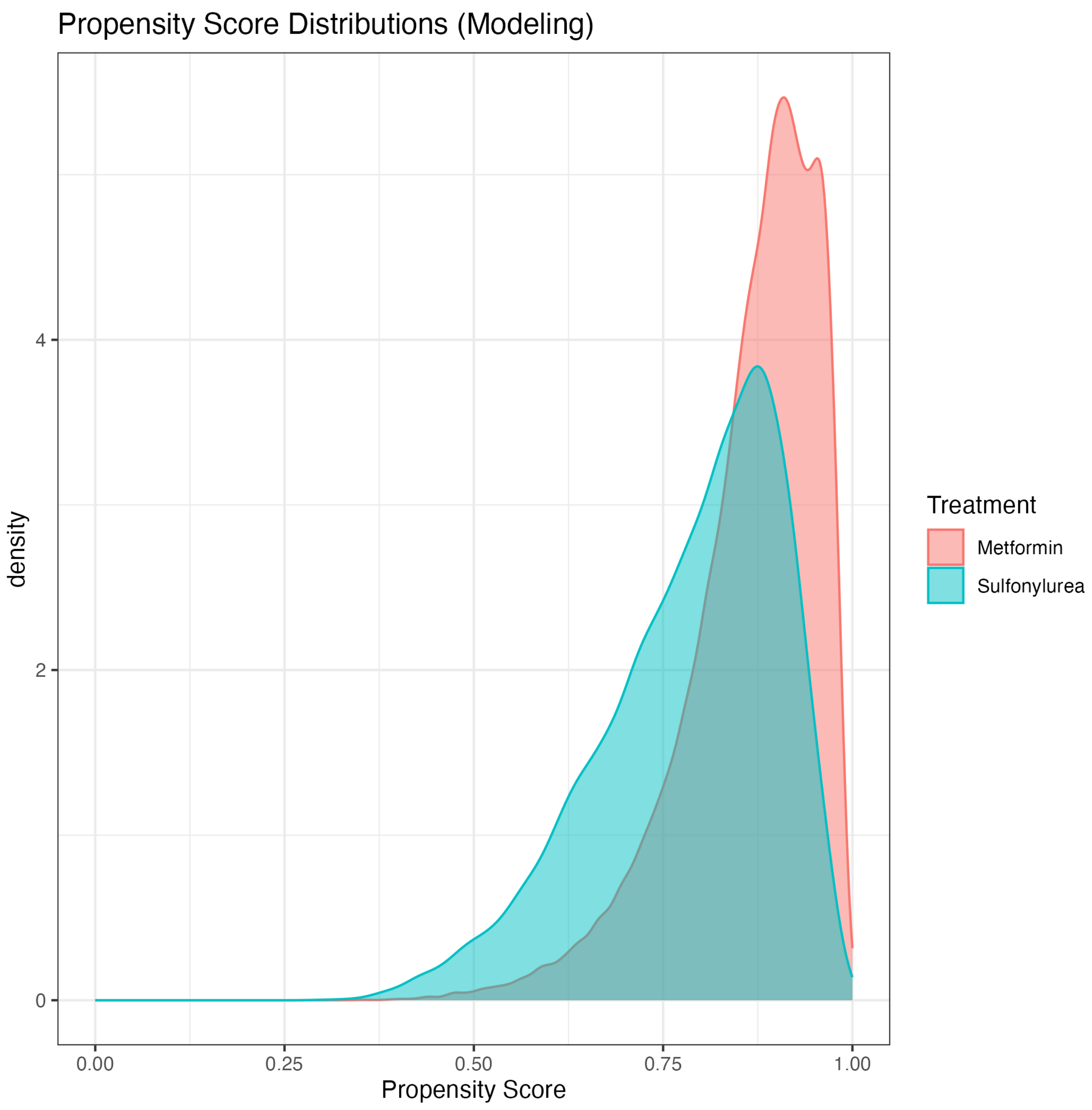


Figure 17: Propensity Distributions (M)

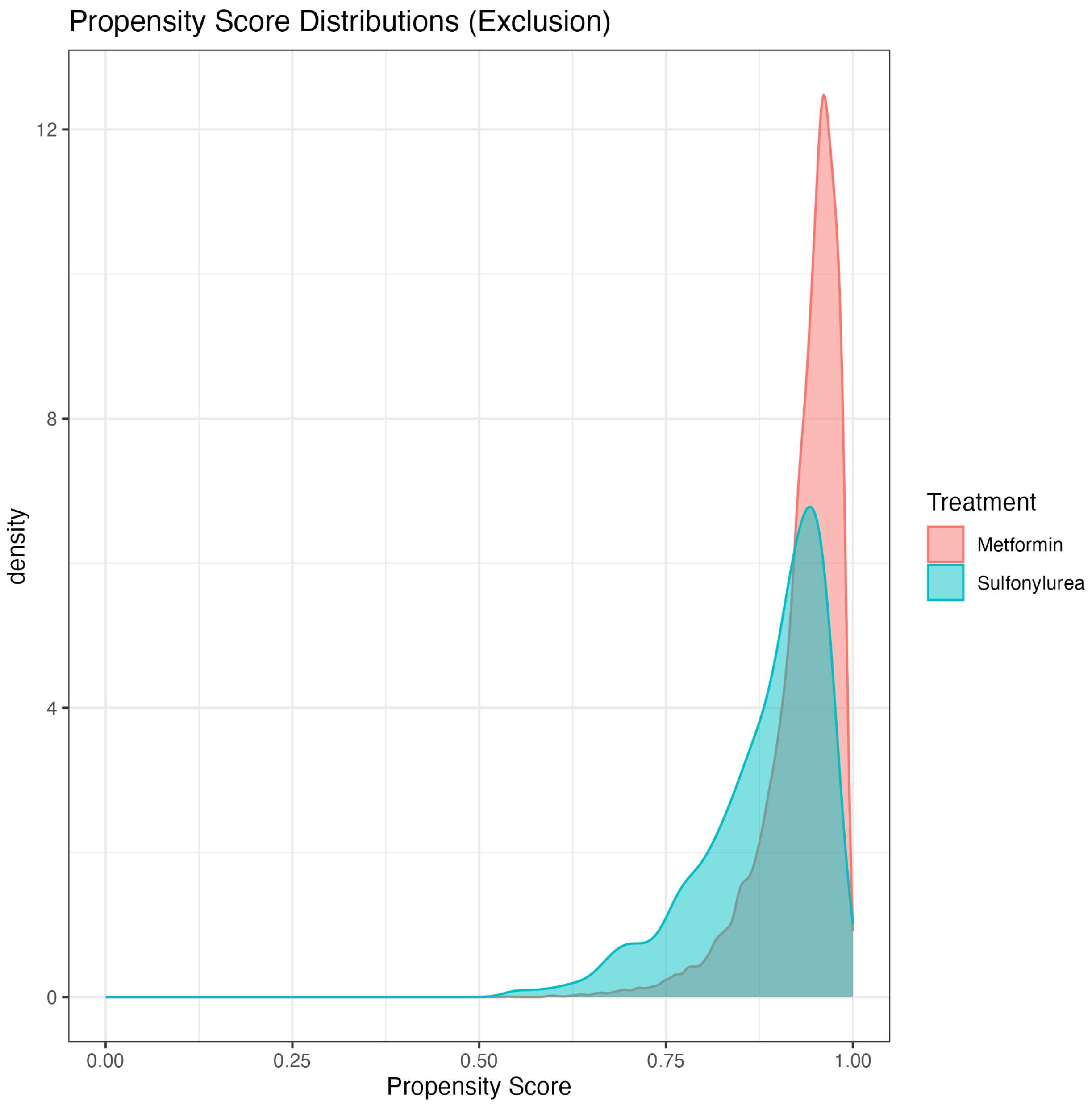


Figure 18: Propensity Distributions (E)

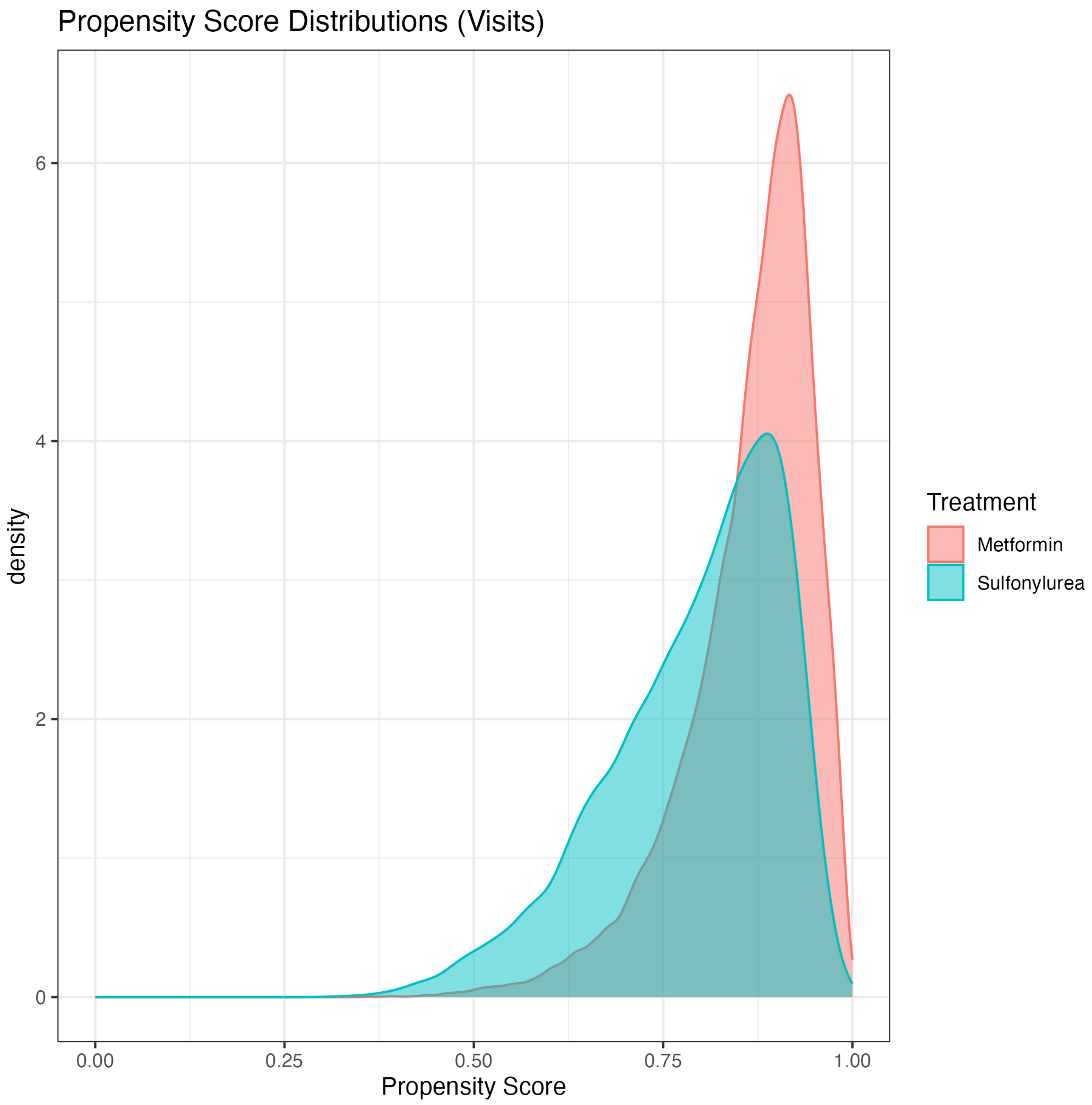


Figure 19: Propensity Distributions (V)

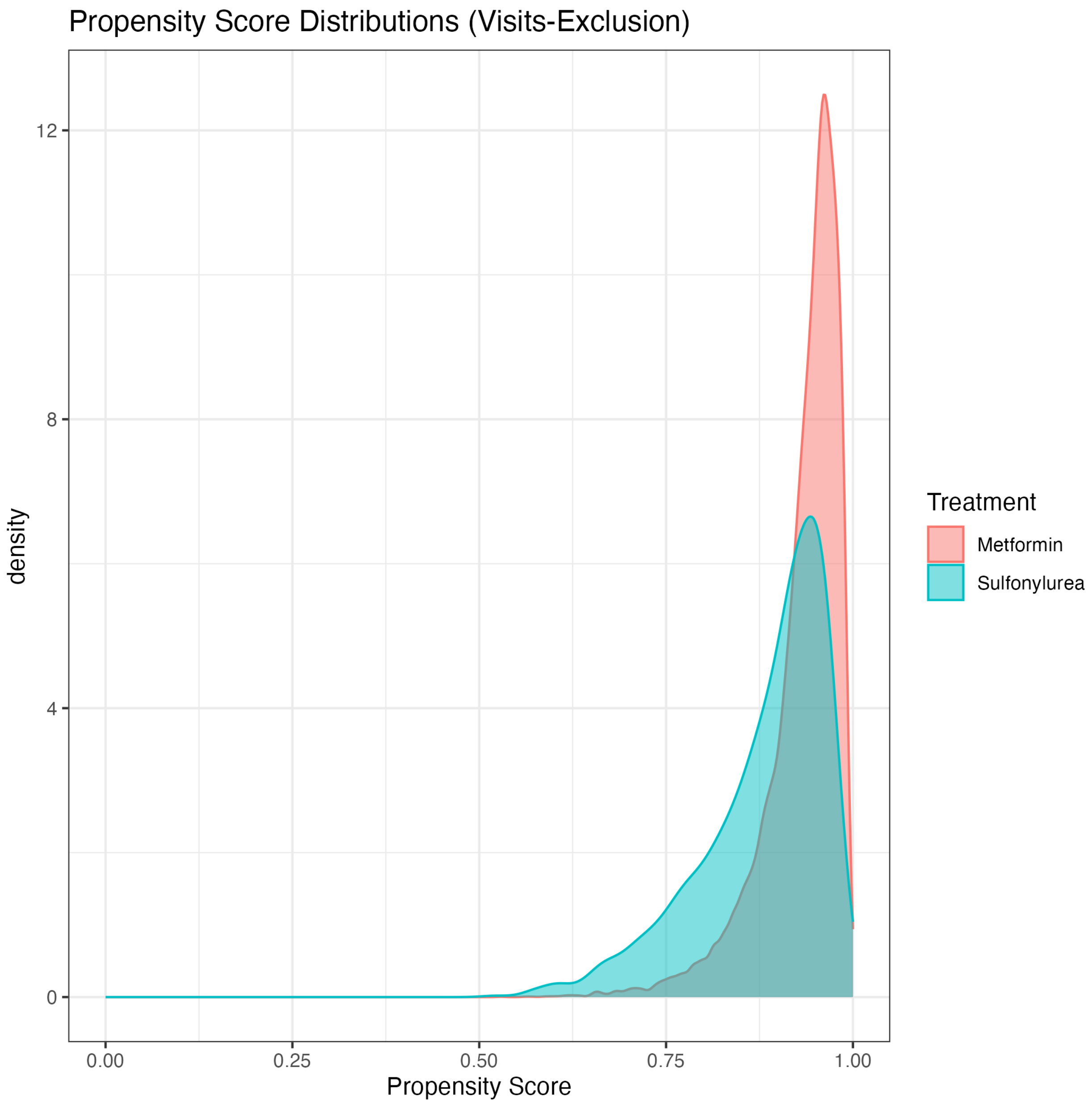


Figure 20: Propensity Distributions (V-E)

### Appendix G. Age-specific Mortality Rates for Alternative Healthcare Criteria

Age-specific mortality rate per 1,000 person-years comparing patients with the top quartile vs. top decile of visits of various types prior to antidiabetic prescription, stratified by sex. Black trend line illustrates the official US census mortality rates reported for MA (Arias, 2022).

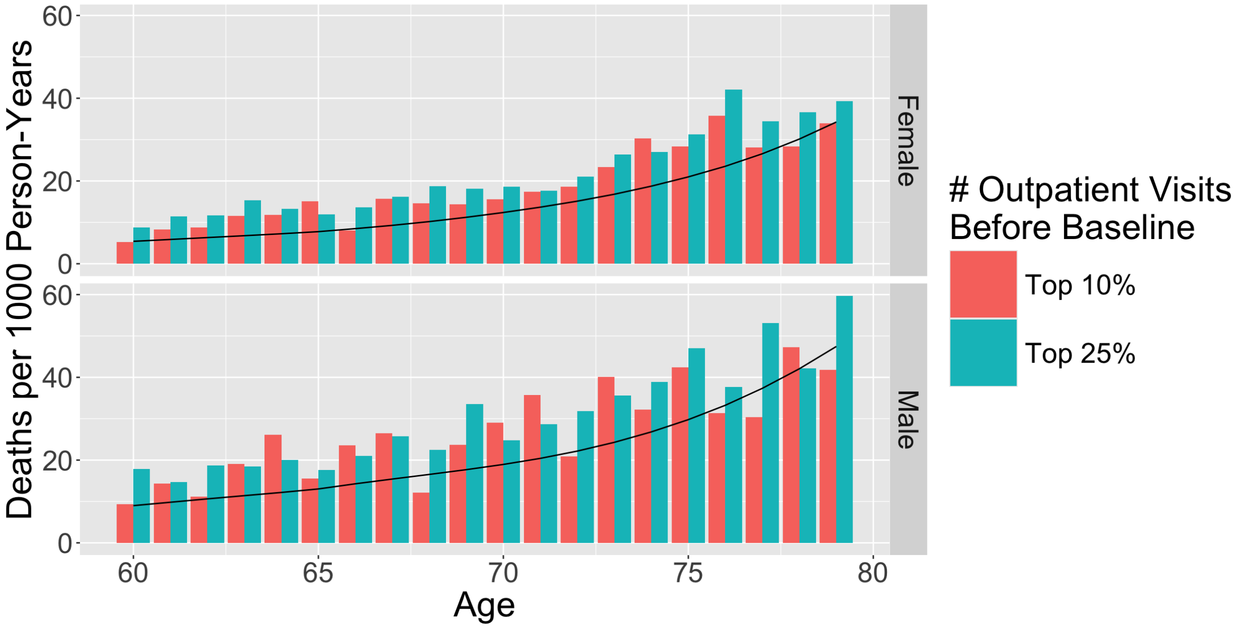


Figure 21: Outpatient Visits (MSE 75%: 1.0, 90%: 1.7)

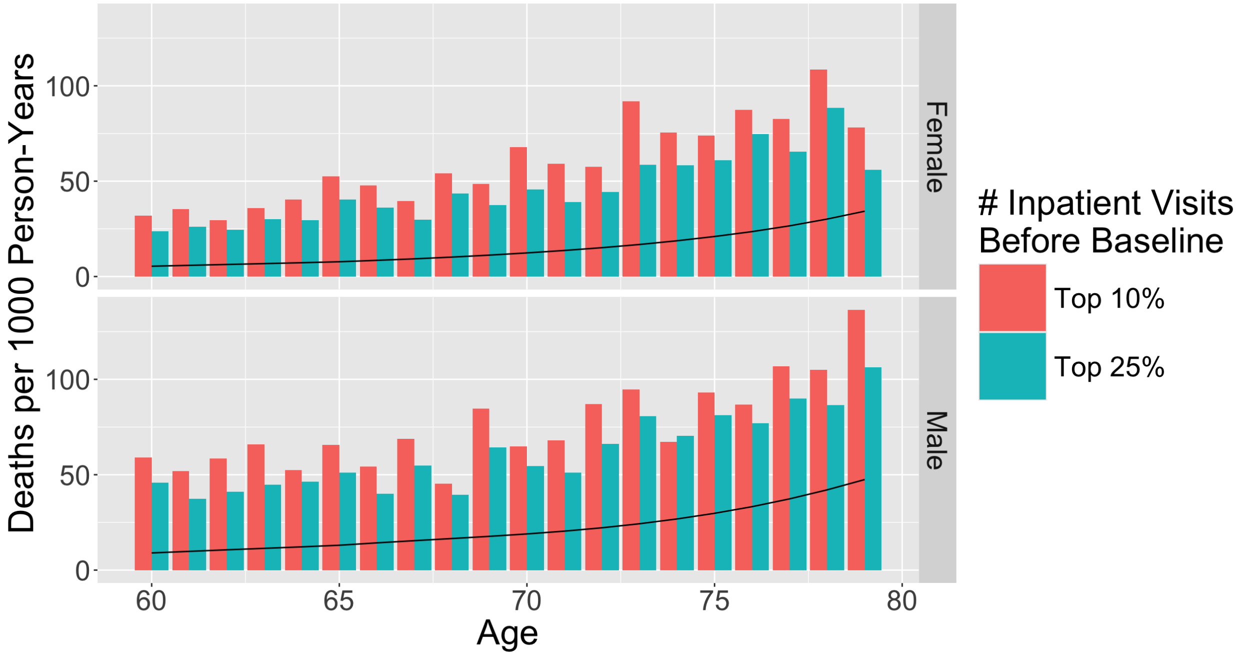


Figure 22: Inpatient Visits (MSE 75%: 30.5, 90%: 15.3)

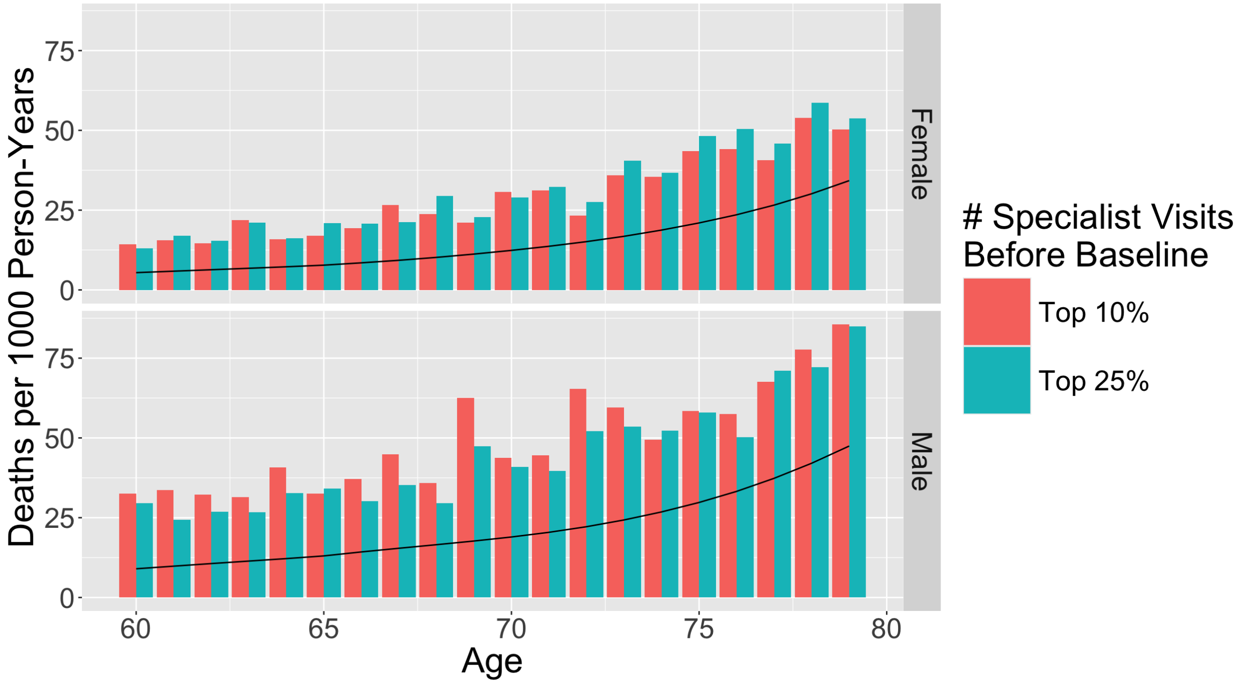


Figure 23: Specialist Visits (MSE 75%: 4.9, 90%: 6.3)

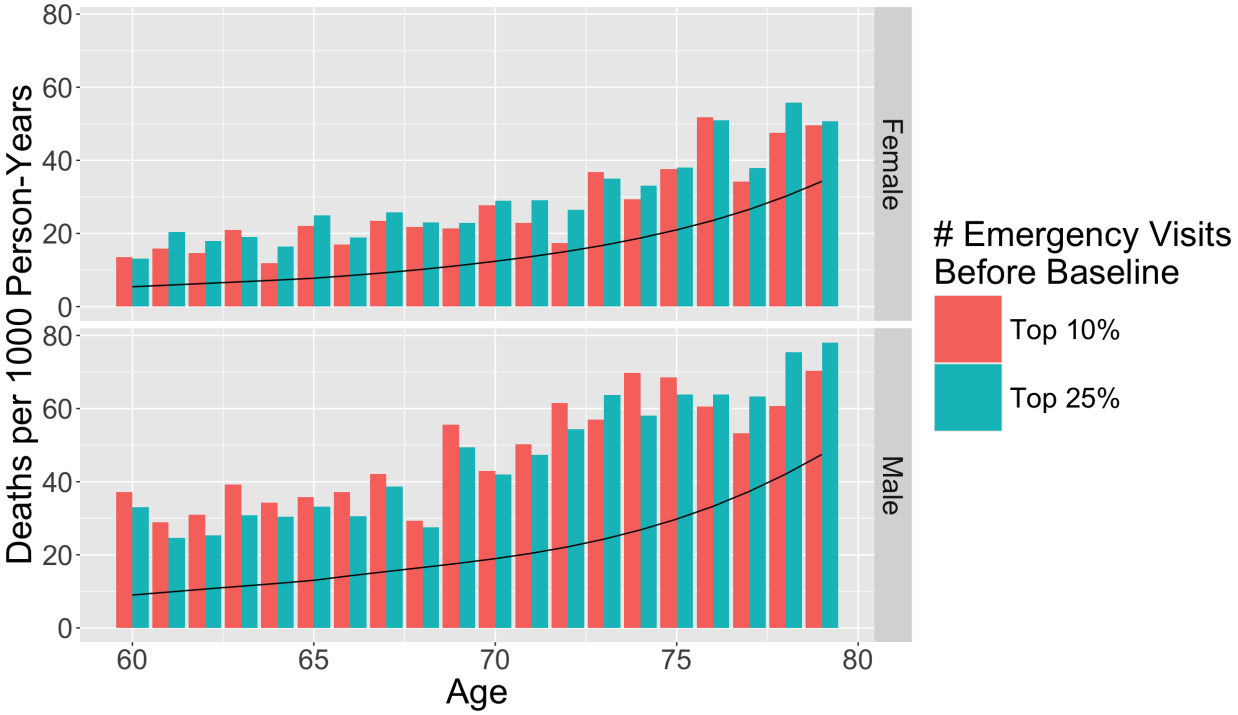


Figure 24: Emergency Visits (MSE 75%: 4.8, 90%: 5.4)

### Appendix H. Age-specific ADRD Diagnosis Rates for Alternative Healthcare Criteria

Age-specific ADRD diagnosis incidence per 1,000 person-years comparing patients with the top quartile vs. top decile of visits of various types prior to antidiabetic prescription. Black trend line is constructed by interpolating data extracted from a figure reported in a study of over eight-million patients using Medicare claims data (Olfson et al., 2021).

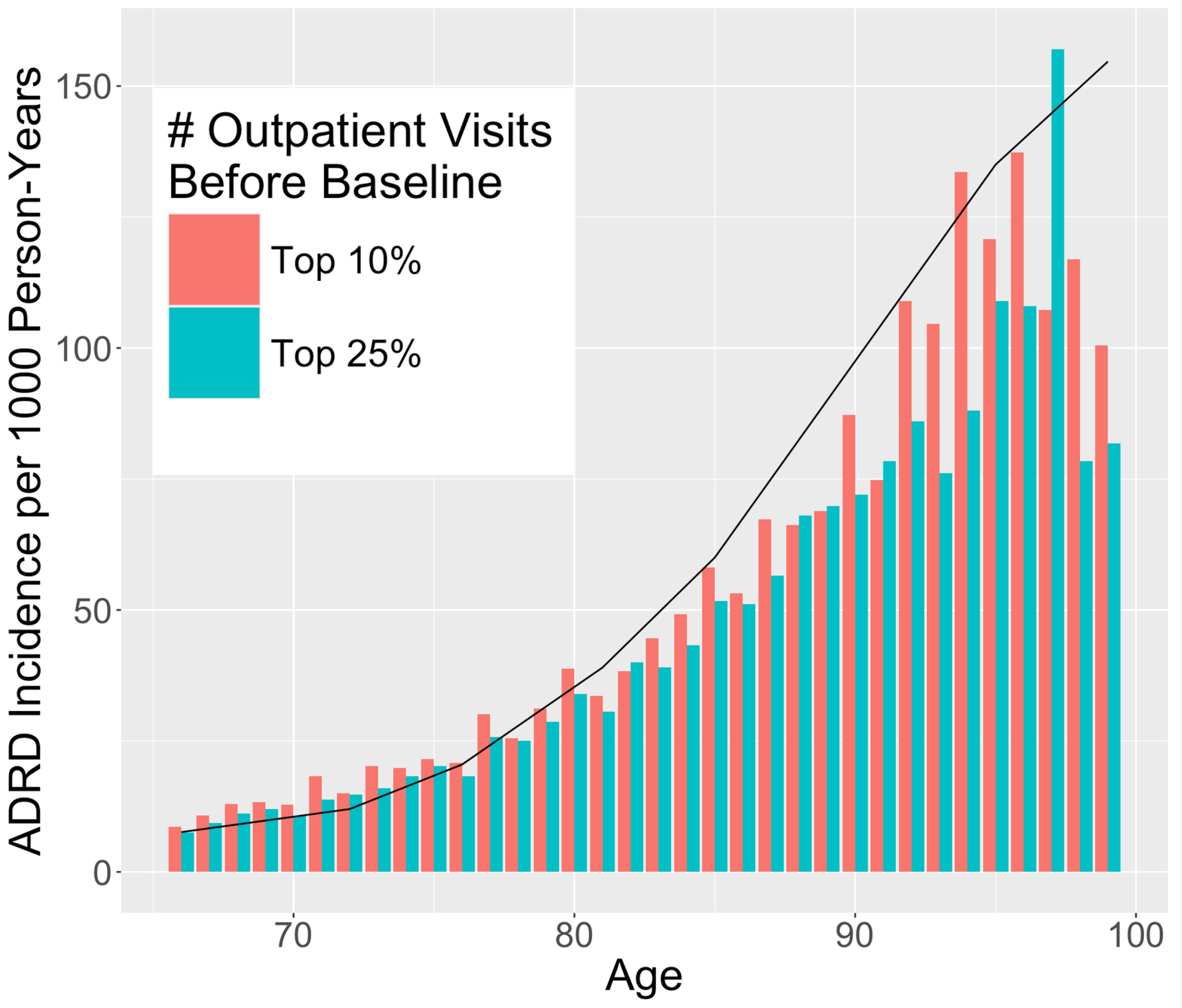


Figure 25: Outpatient Visits (MSE 75%: 1.5, 90%: 0.9)

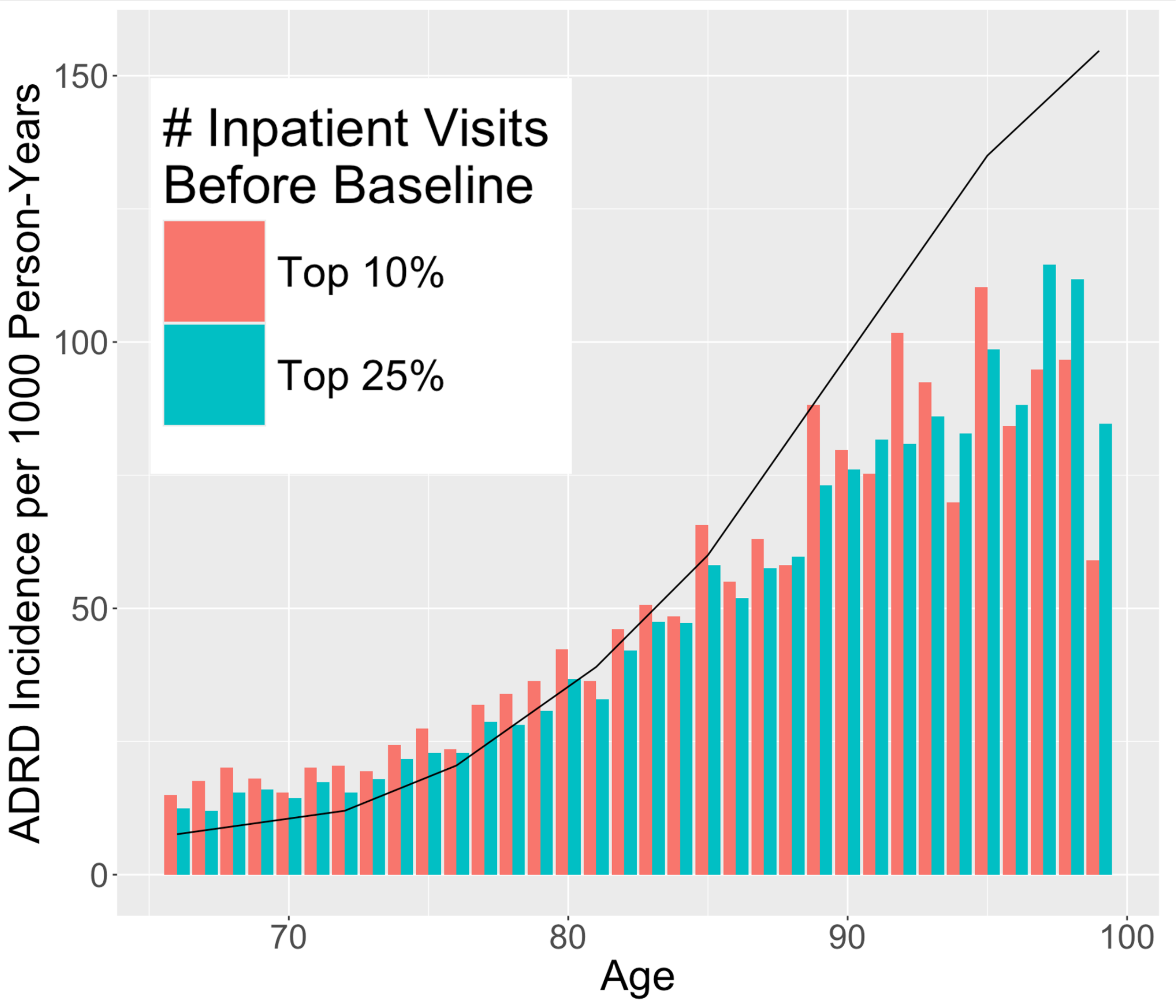


Figure 26: Inpatient Visits (MSE 75%: 2.2, 90%: 3.0)

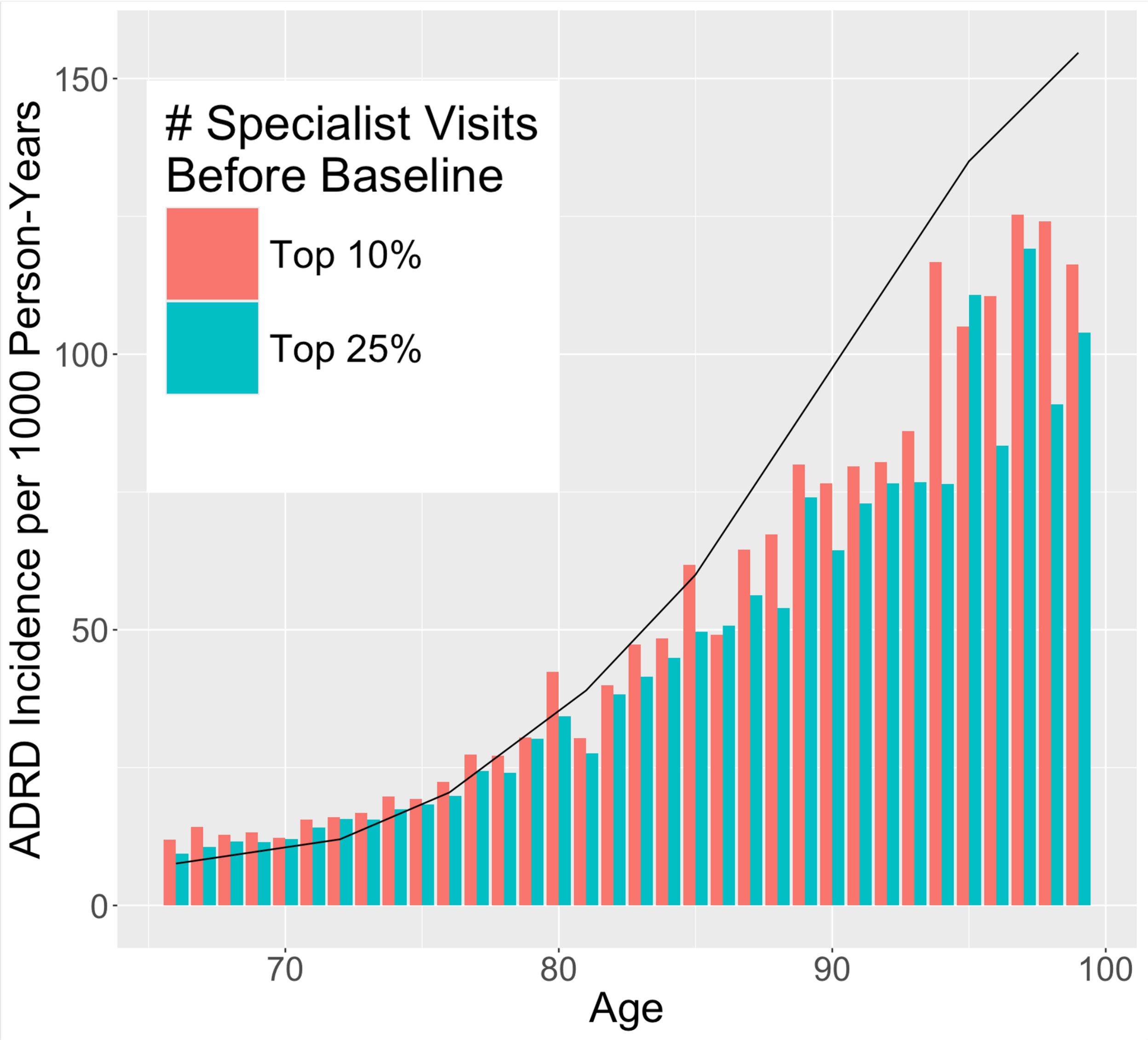


Figure 27: Specialist Visits (MSE 75%: 2.2, 90%: 1.3)

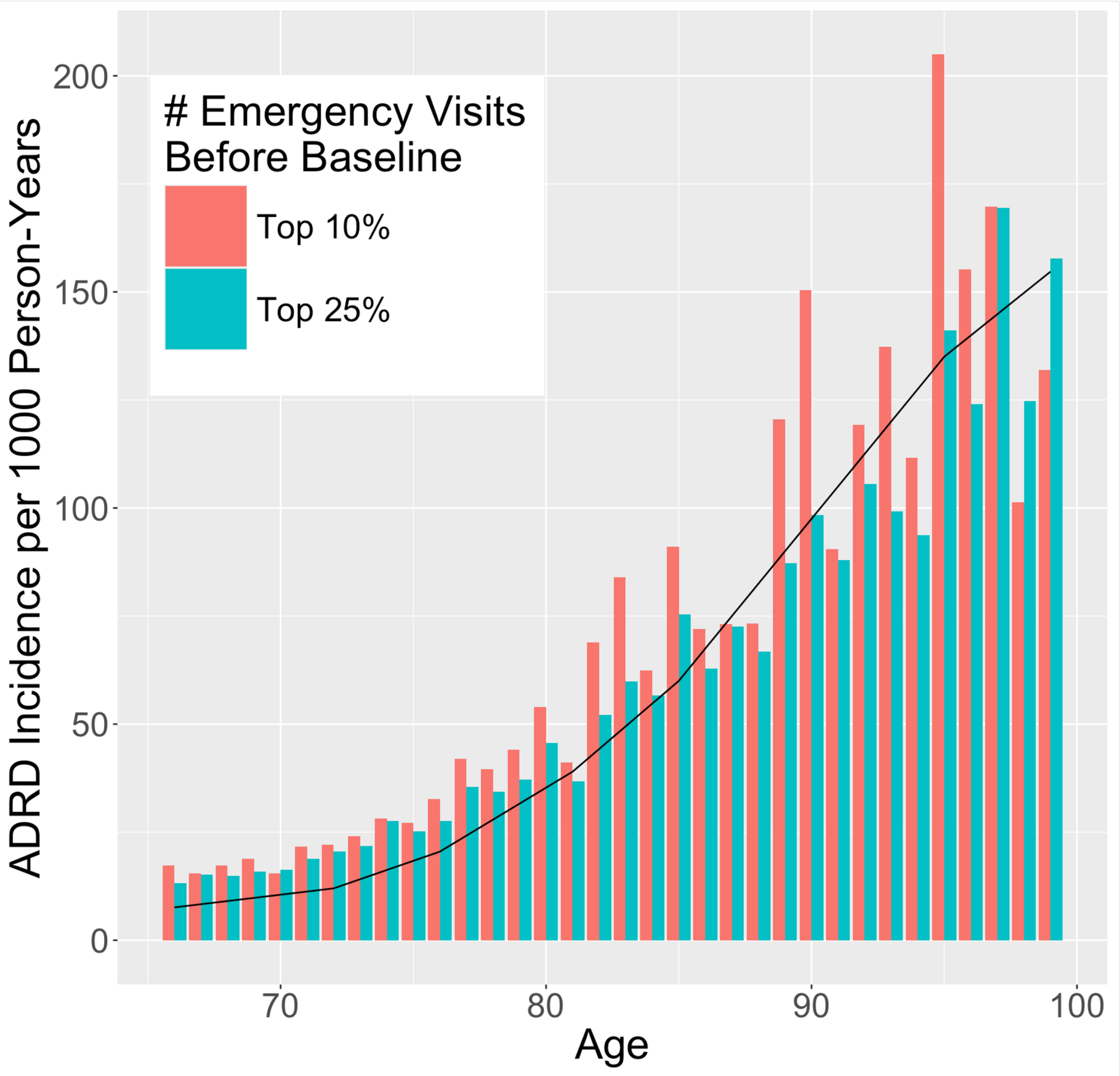


Figure 28: Emergency Visits (MSE 75%: 1.9, 90%: 5.6)

https://www.atsdr.cdc.gov/placeandhealth/svi.

M Olfson, TS Stroup, C Huang, MM Wall, and T Gerhard. Age and incidence of dementia diagnosis. *Journal of General Internal Medicine*, June 2021.

B Vakulenko-Lagun, C Magdamo, ML Charpignon, B Zheng, MW Albers, and S Das. causalcmprsk: An r package for nonparametric and cox-based estimation of average treatment effects in competing risks data. *Computer Methods and Programs in Biomedicine*, 242, 2023.
